# Habitual Coffee Consumption Increases Risks for Metabolic Diseases: Genome-wide Association Studies and a Phenotype-wide Two Sample Mendelian Randomization Analysis

**DOI:** 10.1101/2021.03.08.21253114

**Authors:** Jiuling Li, Tasnim Choudhury, Miaoran Zhang, Lanlan Chen, Jianping Wen, Wanqing Liu, Peng Chen

**Author notes:** Correspondence: Wanqing Liu, PhD. Integrative Biosciences Center, Room 2401, Wayne State University, 6135 Woodward Ave, Detroit, MI 48202, USA., Peng Chen, PhD. College of Basic Medical Sciences, Room 413, Jilin University, 126 Xinmin Street, Changchun, Jilin 130021, China. These authors contributed equally to this work (Wanqing Liu, Peng Chen).

## Abstract

**Background and aims:** Coffee is one of the most widely consumed beverages in the world and has received considerable concerns regarding its impact on human health. Mendelian randomization (MR) could be valuable to explore the potential health effects of coffee via instrumental variables. In this study, we aim to identify novel genetic loci associated with habitual coffee consumption using genome-wide meta-analysis (GWMA) and to evaluate the broad impact of coffee consumption on human health and disease risk via a large-scale, phenotype-wide, two sample Mendelian randomization (TSMR) analysis.

**Methods:** We conducted a genome-wide association study (GWAS) among 283,926 coffee consumers of European ancestry in the UK Biobank (UKBB) to identify single nucleotide polymorphisms (SNPs) associated with the amount of coffee consumption (cups/day, GWAS 1), caffeine intake (GWAS 2) as well as the intake of non-caffeine substance in coffee (GWAS 3). The GWAS 1 results were further combined with the published results from the Coffee and Caffeine Genetics Consortium (CCGC) for a GWMA. TSMR were performed to evaluate the causal-relationship between coffee/caffeine/non-caffeine substance consumption and 1,101 diseases and health traits.

**Results:** The GWMA identified 50 lead SNPs among 19 genomic regions for habitual coffee consumption. Nine out of the 19 loci were novel, including *ADAMTSL4-AS1, CACNA2D2, LINC02123-ADCY2, UBD-SNORD32B, SEMA4D-GADD45G, LOC101929457-LINGO1, RAI1, HCN2,and BRWD1*. The GWAS 2 and 3 identified 2 (*SORCS2* and *SLC39A8*) and 5 (*LINC02060-LINC00461, AGR3-AHR, PRR4-TAS2R14, CYP1A1-CYP1A2*, and *FTO*) genomic regions, respectively. TSMR analysis indicated that coffee consumption increased the risk of high blood lipids, obesity, and diabetes. Meanwhile, intake of caffeine and non-caffeine coffee components decreased and increased some of the blood lipids levels, respectively.

**Conclusions:** Our study provided evidence that habitual coffee consumption could increase the risk of metabolic perturbations. The bioactive components in coffee, other than caffeine, may be more harmful to human health. Our findings have significant implications for global public health given the increasing burden of metabolic diseases.

## Introduction

Coffee is one of the most widely consumed beverages worldwide. The research community has been long debating whether coffee consumption is beneficial or harmful to health. Overall, observational studies favored the beneficial effect of coffee consumption in reducing the risk of metabolic syndrome, obesity, type 2 diabetes (T2D), cardiovascular disease, and several specific cancers^1-4^. However, evidence from randomized controlled trials (RCT) have shed light on the detrimental effects of coffee consumption, such as elevated blood lipids, blood glucose level, and fasting insulin level^5-8^.

The differences in the health outcomes of coffee consumption between the observational studies and RCT highlighted the necessity for further clarification. Mendelian randomization (MR) studies, which were considered to be advantageous to observational studies as they were similar to the RCT design but with comparable length of duration as those of the observational studies. They provided a potentially cost-effective strategy to examine the causal relationship between coffee consumption and health outcomes in human populations. A recent MR study found that coffee consumption increased the risk of osteoarthrosis and obesity^9^, while no significant effect on blood lipids or T2D was identified. Our recent MR analysis did not reveal a significant causal impact of coffee intake on nonalcoholic fatty liver disease (NAFLD)^10^. However, many of these MR studies on coffee consumption were limited in several aspects, with the limited power as a major issue, given the moderate effect of genetic alleles on coffee consumption habit. Therefore, the identification of more genetic susceptibility alleles underlying the coffee drinking habit among larger populations will increase the power for MR analysis. As such, the causal impact of coffee consumption on a broad range of health outcomes can be further elucidated.

In this study, we performed a genome-wide meta-analysis of coffee consumption among 375,388 individuals of European ancestry, which led to the identification of additional novel loci for habitual coffee consumption. Utilizing these expanded list of genetic risk alleles as instrument for coffee consumption, we further conducted two-sample MR (TSMR) analyses between coffee consumption and a large number of health outcomes previously studied among various GWAS. Furthermore, the GWASs of the consumption of caffeine and other coffee-containing, non-caffeine components were also conducted, respectively, and TSMR was also conducted. We observed a significantly detrimental causal impact of habitual coffee consumption and metabolic perturbations, which may be largely attributed to the non-caffeine components in coffee other than caffeine.

## Methods

### Study design and cohorts

The study was conducted using the UK Biobank data resources under application number 53536. As shown in Figure 1, we conducted a GWAS of coffee consumption (GWAS 1) in the UK Biobank cohort (UKBB) and a genome-wide meta-analysis by combining the published results from the Coffee and Caffeine Genetics Consortium (CCGC)^11^. The UKBB cohort included over 500,000 adult individuals recruited from the UK population between 2006 and 2010. The extensive phenotypic and genotypic data were collected among all participants^12^. The quality control on the genotype data followed the procedure recommended by the UKBB^13^. Our GWAS analysis was restricted to the coffee drinkers of European-ancestry. The participants who did not drink coffee were excluded. Finally, 283,926 participants were available for association testing. The genotypic data were further imputed based on the Haplotype Reference Consortium and the UK10K + 1000 genomes reference panel^14^. The CCGC study investigated two phenotypes, the quantitative coffee consumption (phenotype 1) and the comparison between high and low coffee consumers (phenotype 2). In the current study, phenotype 1, which included up to 91,462 coffee consumers of European ancestry, was used in the meta-analysis. This combination resulted in a total sample size of 375,388. Functional annotation of GWAS summary was performed using FUMA GWAS^15^. To investigate the causal effect of coffee consumption, we employed a two-sample MR (TSMR) analysis to screen a broad spectrum of phenotypes in MR-Base^16^. After the enrichment analysis, the significant causal effects were further examined using a one-sample MR (OSMR) analysis using the phenotype defined in UKBB. To distinguish the effects of caffeine and other non-caffeine substances consumption on human health, we carried out the caffeine GWAS (GWAS 2) and other substances GWAS (GWAS 3) using the individual-level data of UKBB. The two phenotypes were defined as below. TSMR was also conducted after FUMA annotation.

**Figure 1.**
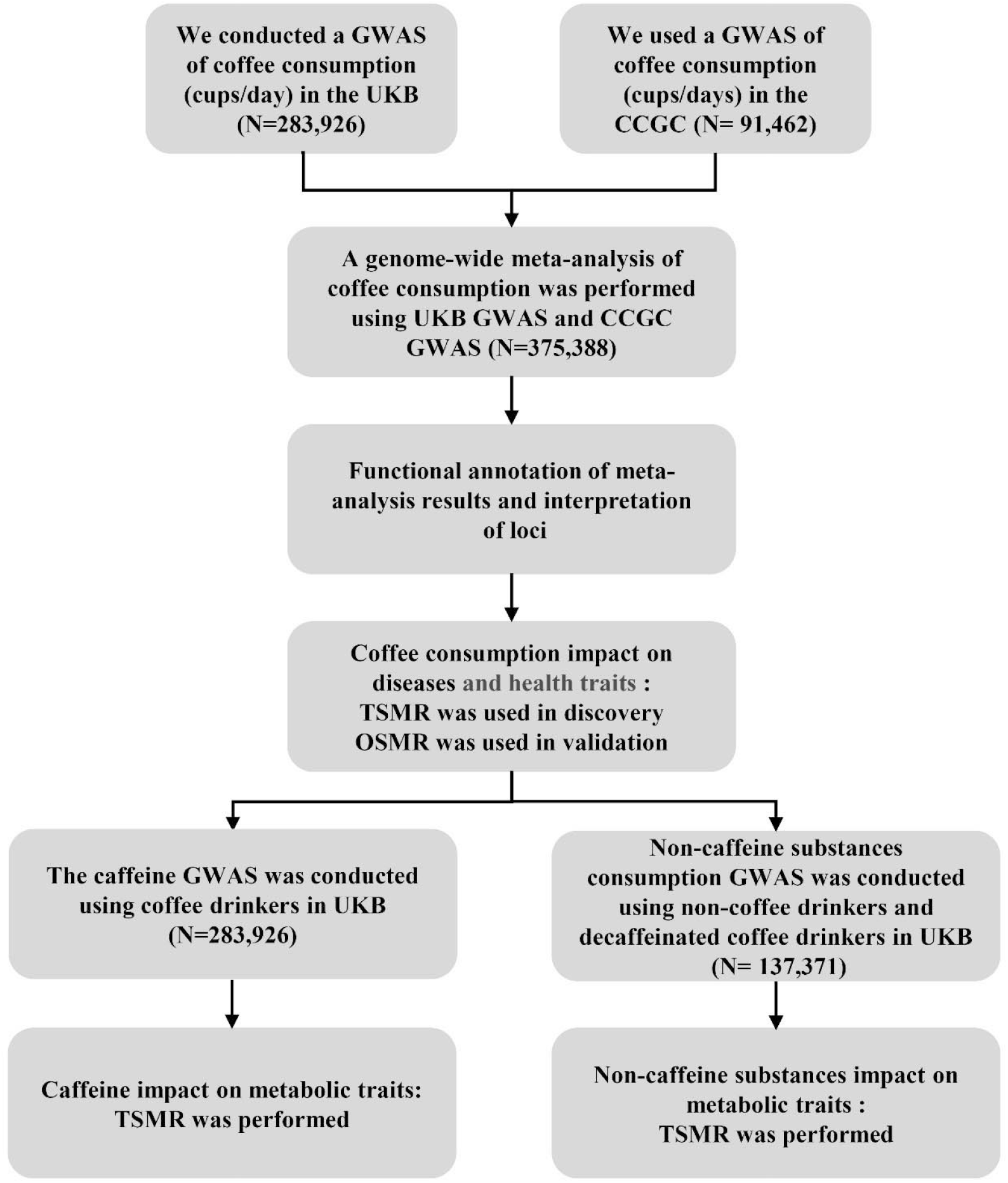
A schematic diagram of the study design.

### Phenotype definitions

Our GWAS 1 was focused on the amount of daily coffee consumption (cups/day). In the UKBB data, coffee consumption (cups/day) and coffee type were surveyed at baseline using the touchscreen questionnaire. The amount of coffee consumption was determined by the question ‘How many cups of coffee do you drink each day (include decaffeinated coffee)?’. If participants reported that they drink less than one cup of coffee per day, their cups per day were set as 0. Participants with very high coffee consumption (> 8 cups/day) were excluded. Notably, the first phenotype of the CCGC study used in the subsequent meta-analysis study was also the number of cups of predominantly regular-type coffee consumed per day among coffee consumers^11^. Therefore, our new GWAS 1 is consistent with this previously conducted GWAS on coffee consumption. The coffee type was also coded and included as a covariate to be adjusted in the GWAS analysis. The coffee type was surveyed by the question “What type of coffee do you usually drink?” The options included decaffeinated coffee, instant coffee, ground coffee and others which were coded as 1, 2, 3, and 4, respectively.

GWAS 2 aimed to identify genetic variants associated with caffeine consumption, in which 283,926 consumers with intake of any type of coffee were included. The consumers of decaffeinated coffee were then coded as category 0, with the consumers of instant coffee, ground coffee, and other types of coffee coded as category 1. GWAS 3 aimed to identify genetic variants associated with other non-caffeine substances contained in coffee consumption, in which “non-coffee drinkers” were coded as 0, with the drinkers consuming decaffeinated coffee as category 1.

### The genome-wide association study and meta-analysis

To identify genetic variants associated with the daily amount of coffee consumption (cups/day), caffeine consumption, and non-caffeine substance consumption, three GWAS were performed using mixed linear model adjusted for age, sex, body mass index, smoking status (never, previous, current), and the first 5 genetic principal components. Coffee type was adjusted for coffee consumption GWAS in addition. The genetic principal components were calculated from the linkage disequilibrium (LD) pruned (r^2^ <0.1) array genotype data of the participants of European ancestry. The autosomal SNPs with minor allele frequency (MAF) > 0.01, imputation INFO score > 0.8, missing rate <0.05, and HWE-Pval >1×10^−6^ were used in the genome-wide association study and meta-analysis. The meta-analysis was performed by combining the GWAS 1 results with that of the CCGC phenotype 1 (cups/day) GWAS using a fixed-effects inverse-variance weighted model^17^.

### Functional annotation of genome-wide association study and meta-analysis

We used the web-based tool FUMA GWAS to define genomic risk loci and obtained functional information of relevant SNPs in these loci^15^. First, lead SNPs were defined using a genome-wide significant P value (5×10^−8^) and LD r^2^ <0.05. All SNPs with significant P value (5×10^−8^) in LD (r^2^ ≥0.05) with one of the lead SNPs were candidate SNPs. Further, genomic risk loci were identified by merging LD blocks if they were less than 250kb apart.

Gene-mapping was based on two strategies. Firstly, positional mapping was performed by selecting exonic and splicing-site SNPs with CADD score ≥12.37^18^. Secondly, expression quantitative trait locus (eQTL) mapping was used to map SNPs to genes that show a significant eQTL association with these SNPs. The eQTL mapping was conducted using data generated in GTEx v8^19^, and only *cis*-eQTLs (SNPs within 1Mb of a gene of interest) were included. The Benjamini-Hochberg false discovery rate (FDR)^20^ of 0.05 was used to define significant eQTL associations. Gene enrichment and tissue specificity expression analysis were conducted using FUMA^15^ and TSEA (http://genetics.wustl.edu/jdlab/tsea/), respectively. We used PhenoScanner to identify the pleiotropic effects of top lead SNPs^21; 22^.

### Mendelian randomization study

The TSMR was performed in an inverse variance weighted (IVW) approach using lead SNPs associated with exposure as instrumental variables (IVs). For coffee consumption as the exposure, we used all 50 lead SNPs reaching the genome-wide level significance as IVs. For both caffeine and non-caffeine coffee intake as exposures, given the small number of SNPs reaching the genome-wide significance level, we used SNPs with a p value <10^−5^ as IVs to reduce potential pleotropic effects. This results 38 and 83 IVs for caffeine and non-caffeine substance intake, respectively. For all candidate IVs, only when the p-value of the IVs in an outcome is at least greater than 0.001, can it be used to infer the causality between the exposure and outcomes. The outcomes were the phenotypes available in MR-Base (N=1,101). MR-Egger regression and Cochran’s Q test were used to detect the pleiotropic effect and the heterogeneity of the IVs. The causal effects estimated using the IVs with MR-egger intercept p value ≤0.05 or Cochran’s Q p value ≤0.05 were considered to be biased. The enrichment analysis of the significant causal effects (IVW p ≤0.05) in the categories defined by MR-Base was conducted using hypergeometric distribution test (https://systems.crump.ucla.edu/hypergeometric/index.php).

To reduce the false positive rate, the significant causal effects of coffee consumption (cups/day) on health outcomes were further validated using OSMR analysis using individual level UKBB data. In our OSMR study, the causal effect of coffee consumption on an outcome was estimated by the association between the coffee polygenic risk score (coffee-PRS) and the outcome. For each participant in the UKBB, the coffee PRS was calculated by adding together the allele dosages of the instrumental variables, weighted by their association effects with coffee consumption. The association with a dichotomous or continuous outcome was estimated using a logistic regression or linear regression model, respectively, without an adjustment. For random blood glucose, the linear model was adjusted for the self-report fasting time.

### Statistical analysis

The linear mixed model was estimated using the Genome-wide Complex Trait Analysis (GCTA)^23^. The genome-wide meta-analysis was performed using METAL^17^. The TSMR analysis was conducted using the TwoSampleMR package of R (version 0.4.25). The polygenic risk score (PRS) and the explanation of the coffee consumption by IVs (r^2^) were evaluated by PRSice-2^24^. The F-statistic was calculated by the following formula to estimate the statistical power of lead SNPs:

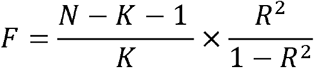

N was the sample size, and k was the number of IVs.

Linear regression and logistic regression were conducted using R software (version 4.0.2, https://www.r-project.org/).

## Results

### GWAS of coffee consumption (cups/day), caffeine consumption, and non-caffeine coffee consumption

Our GWAS 1 on coffee consumption involved 283,926 coffee consumers and 9,462,639 SNPs with MAF>0.01, imputation INFO score>0.8, and missing rate <0.05. The full details of the samples are provided in Supplementary Table 1. At this stage, we found 18 loci (Supplementary Table 2). After the meta-analysis by further combining the data of the CCGC GWAS, we were able to identify an additional significant locus (rs1571536, *SEMA4D-GADD45G*) (Figure 2A, Supplementary Table 3). Regional plots are available in the online resources (Supplementary Figure S1-2). Of the total of 19 identified loci, 6 loci including rs1260326 (*GCKR*), rs1481012 (*ABCG2*), rs4410790 (*AGR3-AHR*), rs799166 (*MLXIPL-VPS37D*), rs17685 (*POR*), and rs2472297 (*CYP1A1-CYP1A2*), were previously reported by CCGC^11^. Four loci, rs2867110 (*LOC105373352-TMEM18*), rs476828 (*PMAIP1-MC4R*), rs56113850 (*CYP2A6)*, and rs6512309 (*PCMTD2*), were identified by Zhong VW et al^25^. The 9 newly identified loci were rs6655975 (*ADAMTSL4-AS1*), rs1467913 *(CACNA2D2)*, rs12519880*(LINC02123-ADCY2)*, rs1235162*(UBD-SNORD32B)*, rs1571536(*SEMA4D-GADD45G*), rs2667773 (*LOC101929457-LINGO1*), rs11078398 *(RAI1)*, rs113534512 *(HCN2)*, and rs3945 (*BRWD1*). Among which, rs6655975 (*ADAMTSL4-AS1*), rs1571536 (*SEMA4D-GADD45G*), rs2667773 (*LOC101929457-LINGO1*), and rs3945 (*BRWD1*) were nominally significant in the CCGC GWAS study at a significant level of 0.05 with the same direction for the associations (Table 1). For the locus 9 (*MLXIPL-VPS37D*), rs7800944 was identified as an index SNP in CCGC. In our results, the lead SNP rs799166 is in LD (r^2^ =0.36, among European Caucasian population) with rs7800944, and was predicted to be located in a SMAD2 binding site in JASPAR^26^. For the loci 13 and 17, the lead SNPs (rs2667773 and rs56113850) were not present in the CCGC GWAS findings. However, their LD proxy SNPs (rs2667768 and rs1496402) (r^2^ >0.6 with the aforementioned lead SNPs among Caucasian population) were observed as the corresponding lead SNPs, respectively. Therefore, of the 50 lead SNPs, 35 were nominally validated in the previous CCGC study (p<0.05), while 15 were newly observed as lead SNPs only in the current meta-analysis (Supplementary Table 4).

**Figure 2.**
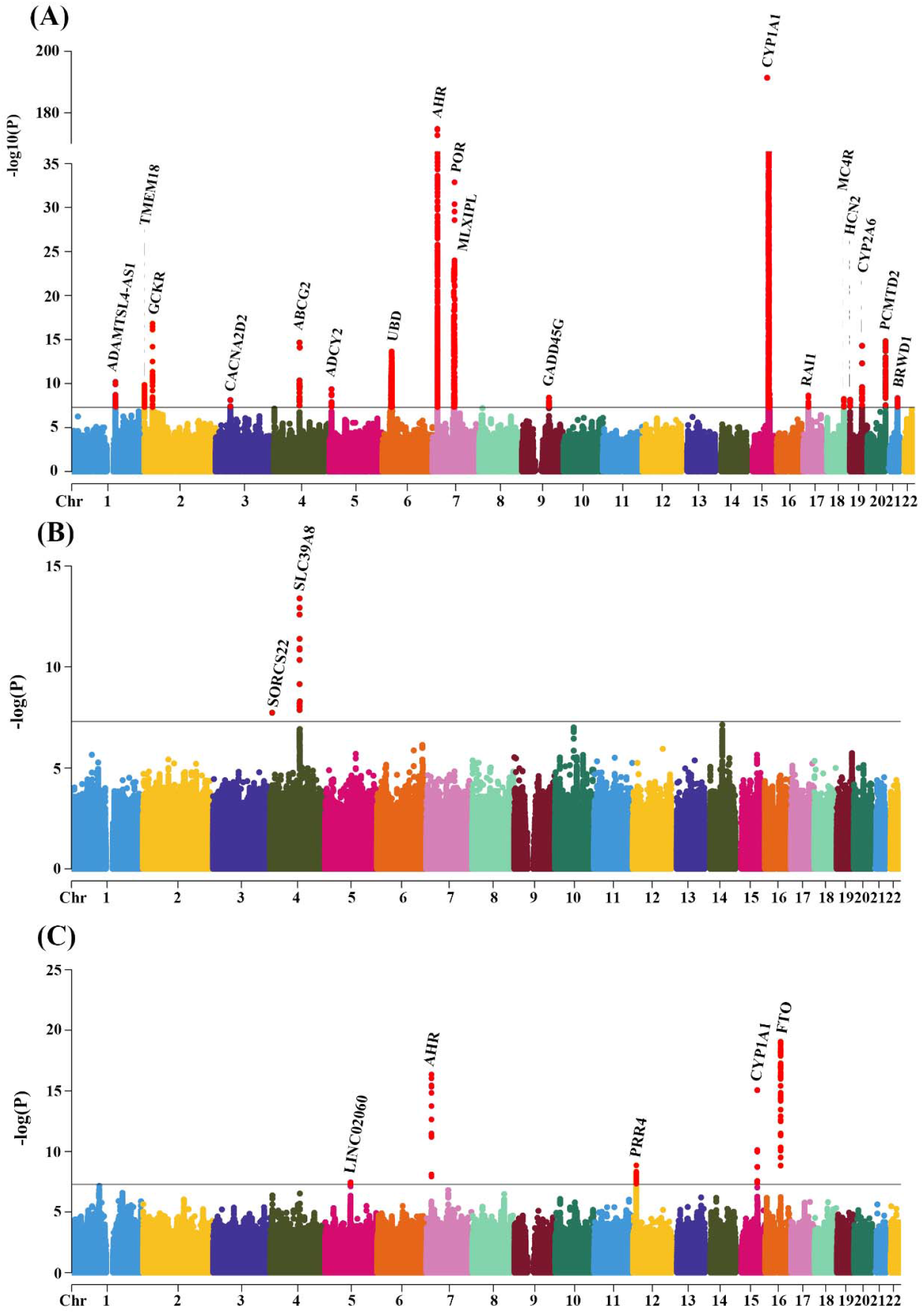
The Manhattan plot displays the genome-wide associations between SNPs and coffee consumption (A), caffeine consumption (B), and non-caffeine substances consumption (C). The x-axis represents genomic position of variants. The y-axis shows the strength of the associations (–log_10_ P). The dash line indicates the genome wide significance level of p=5e-8

**Table 1.**
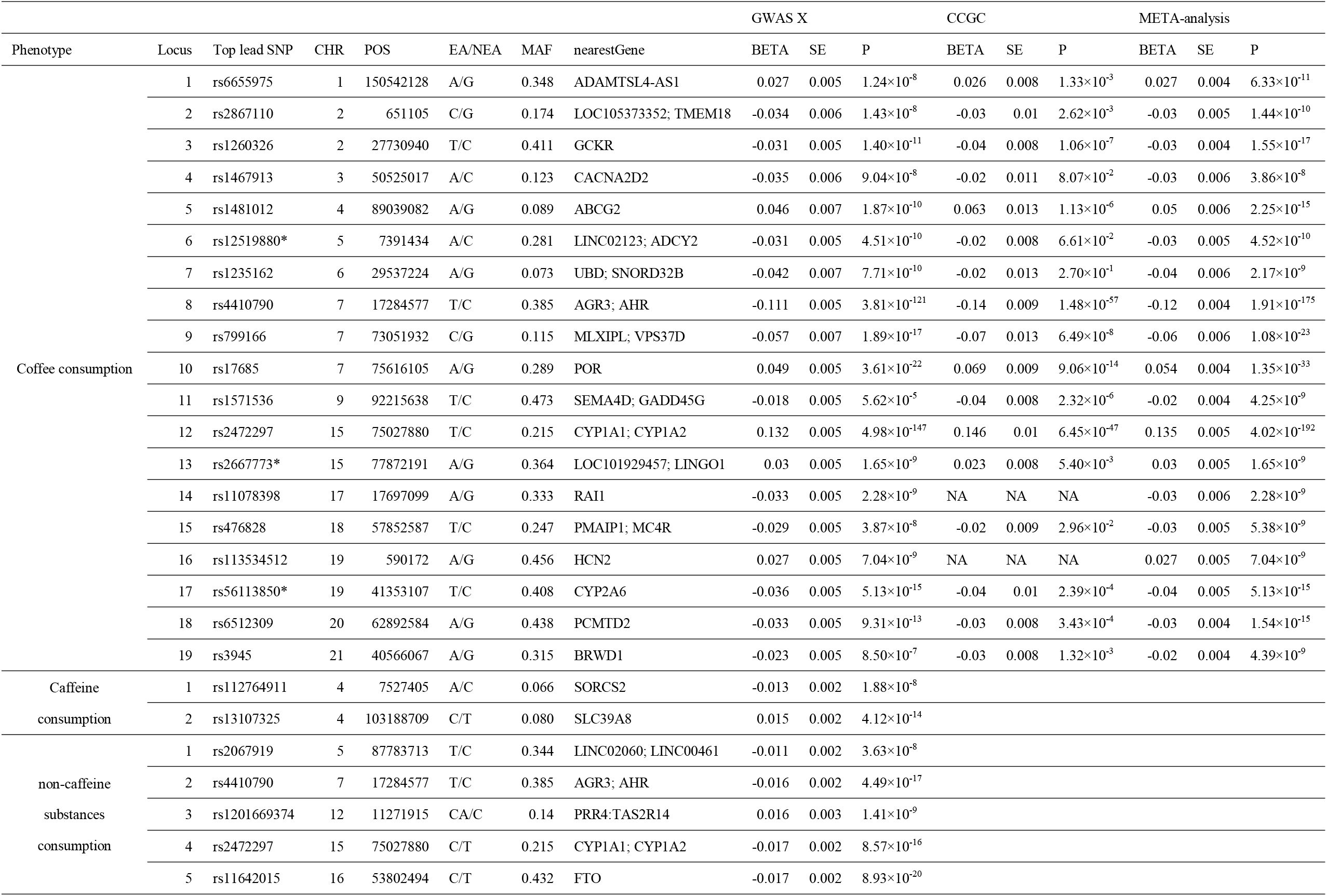
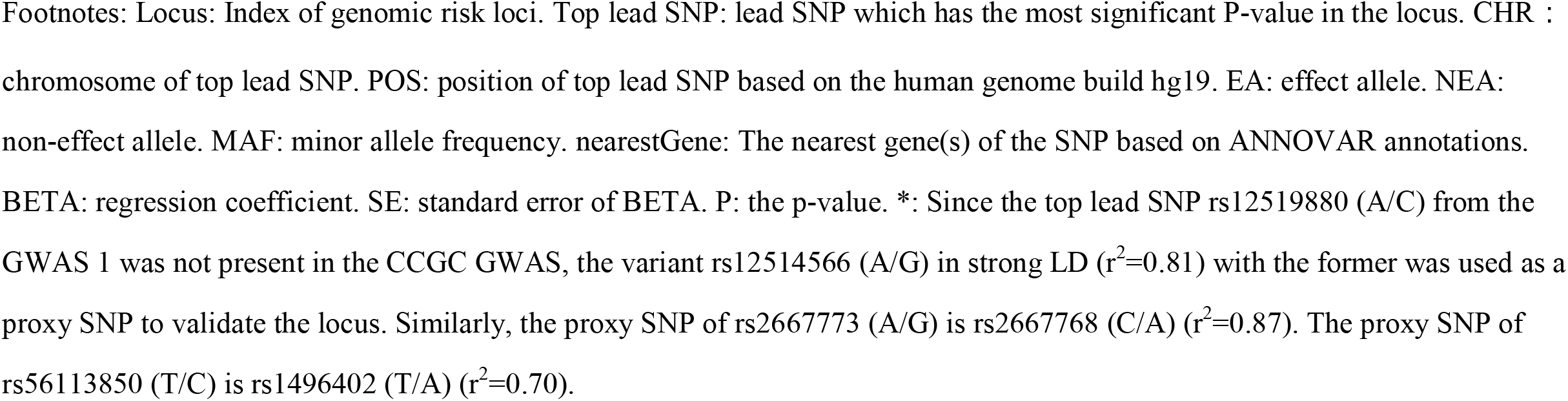
The Top SNPs significantly associated with coffee consumption, caffeine consumption, and non-caffeine substances consumption, respectively.

| Phenotype | Locus | Top lead SNP | CHR | POS | EA/NEA | MAF | nearestGene | GWAS X |  |  | CCGC |  |  | META-analysis |  |  |
| --- | --- | --- | --- | --- | --- | --- | --- | --- | --- | --- | --- | --- | --- | --- | --- | --- |
|  |  |  |  |  |  |  |  | BETA | SE | P | BETA | SE | P | BETA | SE | P |
| Coffee consumption | 1 | rs6655975 | 1 | 150542128 | A/G | 0.348 | ADAMTSL4-AS1 | 0.027 | 0.005 | 1.24×10<sup>-8</sup> | 0.026 | 0.008 | 1.33×10<sup>-3</sup> | 0.027 | 0.004 | 6.33×10<sup>-11</sup> |
|  | 2 | rs2867110 | 2 | 651105 | C/G | 0.174 | LOC105373352; TMEM18 | -0.034 | 0.006 | 1.43×10<sup>-8</sup> | -0.03 | 0.01 | 2.62×10<sup>-3</sup> | -0.03 | 0.005 | 1.44×10<sup>-10</sup> |
|  | 3 | rs1260326 | 2 | 27730940 | T/C | 0.411 | GCKR | -0.031 | 0.005 | 1.40×10<sup>-11</sup> | -0.04 | 0.008 | 1.06×10<sup>-7</sup> | -0.03 | 0.004 | 1.55×10<sup>-17</sup> |
|  | 4 | rs1467913 | 3 | 50525017 | A/C | 0.123 | CACNA2D2 | -0.035 | 0.006 | 9.04×10<sup>-8</sup> | -0.02 | 0.011 | 8.07×10<sup>-2</sup> | -0.03 | 0.006 | 3.86×10<sup>-8</sup> |
|  | 5 | rs1481012 | 4 | 89039082 | A/G | 0.089 | ABCG2 | 0.046 | 0.007 | 1.87×10<sup>-10</sup> | 0.063 | 0.013 | 1.13×10<sup>-6</sup> | 0.05 | 0.006 | 2.25×10<sup>-15</sup> |
|  | 6 | rs12519880* | 5 | 7391434 | A/C | 0.281 | LINC02123; ADCY2 | -0.031 | 0.005 | 4.51×10<sup>-10</sup> | -0.02 | 0.008 | 6.61×10<sup>-2</sup> | -0.03 | 0.005 | 4.52×10<sup>-10</sup> |
|  | 7 | rs1235162 | 6 | 29537224 | A/G | 0.073 | UBD; SNORD32B | -0.042 | 0.007 | 7.71×10<sup>-10</sup> | -0.02 | 0.013 | 2.70×10<sup>-1</sup> | -0.04 | 0.006 | 2.17×10<sup>-9</sup> |
|  | 8 | rs4410790 | 7 | 17284577 | T/C | 0.385 | AGR3; AHR | -0.111 | 0.005 | 3.81×10<sup>-121</sup> | -0.14 | 0.009 | 1.48×10<sup>-57</sup> | -0.12 | 0.004 | 1.91×10<sup>-175</sup> |
|  | 9 | rs799166 | 7 | 73051932 | C/G | 0.115 | MLXIPL; VPS37D | -0.057 | 0.007 | 1.89×10<sup>-17</sup> | -0.07 | 0.013 | 6.49×10<sup>-8</sup> | -0.06 | 0.006 | 1.08×10<sup>-23</sup> |
|  | 10 | rs17685 | 7 | 75616105 | A/G | 0.289 | POR | 0.049 | 0.005 | 3.61×10<sup>-22</sup> | 0.069 | 0.009 | 9.06×10<sup>-14</sup> | 0.054 | 0.004 | 1.35×10<sup>-33</sup> |
|  | 11 | rs1571536 | 9 | 92215638 | T/C | 0.473 | SEMA4D; GADD45G | -0.018 | 0.005 | 5.62×10<sup>-5</sup> | -0.04 | 0.008 | 2.32×10<sup>-6</sup> | -0.02 | 0.004 | 4.25×10<sup>-9</sup> |
|  | 12 | rs2472297 | 15 | 75027880 | T/C | 0.215 | CYP1A1; CYP1A2 | 0.132 | 0.005 | 4.98×10<sup>-147</sup> | 0.146 | 0.01 | 6.45×10<sup>-47</sup> | 0.135 | 0.005 | 4.02×10<sup>-192</sup> |
|  | 13 | rs2667773* | 15 | 77872191 | A/G | 0.364 | LOC101929457; LINGO1 | 0.03 | 0.005 | 1.65×10<sup>-9</sup> | 0.023 | 0.008 | 5.40×10<sup>-3</sup> | 0.03 | 0.005 | 1.65×10<sup>-9</sup> |
|  | 14 | rs11078398 | 17 | 17697099 | A/G | 0.333 | RAI1 | -0.033 | 0.005 | 2.28×10<sup>-9</sup> | NA | NA | NA | -0.03 | 0.006 | 2.28×10<sup>-9</sup> |
|  | 15 | rs476828 | 18 | 57852587 | T/C | 0.247 | PMAIP1; MC4R | -0.029 | 0.005 | 3.87×10<sup>-8</sup> | -0.02 | 0.009 | 2.96×10<sup>-2</sup> | -0.03 | 0.005 | 5.38×10<sup>-9</sup> |
|  | 16 | rs113534512 | 19 | 590172 | A/G | 0.456 | HCN2 | 0.027 | 0.005 | 7.04×10<sup>-9</sup> | NA | NA | NA | 0.027 | 0.005 | 7.04×10<sup>-9</sup> |
|  | 17 | rs56113850* | 19 | 41353107 | T/C | 0.408 | CYP2A6 | -0.036 | 0.005 | 5.13×10<sup>-15</sup> | -0.04 | 0.01 | 2.39×10<sup>-4</sup> | -0.04 | 0.005 | 5.13×10<sup>-15</sup> |
|  | 18 | rs6512309 | 20 | 62892584 | A/G | 0.438 | PCMTD2 | -0.033 | 0.005 | 9.31×10<sup>-13</sup> | -0.03 | 0.008 | 3.43×10<sup>-4</sup> | -0.03 | 0.004 | 1.54×10<sup>-15</sup> |
|  | 19 | rs3945 | 21 | 40566067 | A/G | 0.315 | BRWD1 | -0.023 | 0.005 | 8.50×10<sup>-7</sup> | -0.03 | 0.008 | 1.32×10<sup>-3</sup> | -0.02 | 0.004 | 4.39×10<sup>-9</sup> |
| Caffeine consumption | 1 | rs112764911 | 4 | 7527405 | A/C | 0.066 | SORCS2 | -0.013 | 0.002 | 1.88×10<sup>-8</sup> |  |  |  |  |  |  |
|  | 2 | rs13107325 | 4 | 103188709 | C/T | 0.080 | SLC39A8 | 0.015 | 0.002 | 4.12×10<sup>-14</sup> |  |  |  |  |  |  |
| non-caffeine substances consumption | 1 | rs2067919 | 5 | 87783713 | T/C | 0.344 | LINC02060; LINC00461 | -0.011 | 0.002 | 3.63×10<sup>-8</sup> |  |  |  |  |  |  |
|  | 2 | rs4410790 | 7 | 17284577 | T/C | 0.385 | AGR3; AHR | -0.016 | 0.002 | 4.49×10<sup>-17</sup> |  |  |  |  |  |  |
|  | 3 | rs1201669374 | 12 | 11271915 | CA/C | 0.14 | PRR4; TAS2R14 | 0.016 | 0.003 | 1.41×10<sup>-9</sup> |  |  |  |  |  |  |
|  | 4 | rs2472297 | 15 | 75027880 | C/T | 0.215 | CYP1A1; CYP1A2 | -0.017 | 0.002 | 8.57×10<sup>-16</sup> |  |  |  |  |  |  |
|  | 5 | rs11642015 | 16 | 53802494 | C/T | 0.432 | FTO | -0.017 | 0.002 | 8.93×10<sup>-20</sup> |  |  |  |  |  |  |

Footnotes: Locus: Index of genomic risk loci. Top lead SNP: lead SNP which has the most significant P-value in the locus. CHR : chromosome of top lead SNP. POS: position of top lead SNP based on the human genome build hg19. EA: effect allele. NEA: non-effect allele. MAF: minor allele frequency. nearestGene: The nearest gene(s) of the SNP based on ANNOVAR annotations. BETA: regression coefficient. SE: standard error of BETA. P: the p-value. \*: Since the top lead SNP rs12519880 (A/C) from the GWAS 1 was not present in the CCGC GWAS, the variant rs12514566 (A/G) in strong LD ( $r^2=0.81$ ) with the former was used as a proxy SNP to validate the locus. Similarly, the proxy SNP of rs2667773 (A/G) is rs2667768 (C/A) ( $r^2=0.87$ ). The proxy SNP of rs56113850 (T/C) is rs1496402 (T/A) ( $r^2=0.70$ ).

The participants and SNPs used in the GWAS 2 were the same as the GWAS 1. At this stage, we found rs112764911 (*SORCS2*) and rs13107325 (*SLC39A8*) to be associated with caffeine consumption at the genome-wide level (Table 1, Figure 2B). Regional plots are available in the online resources (Supplementary Figure S3-4).

Our GWAS 3 of non-caffeine substances consumption involved 137,371 participants and 9,462,277 SNPs with MAF>0.01, imputation INFO score>0.8, and missing rate <0.05. We identified 5 lead SNPs associated with non-caffeine substances consumption, including rs2067919 (*LINC02060*), rs4410790 (*AHR*), rs1201669374 (*PRR4*), rs2472297 (*CYP1A1*), and rs11642015 (*FTO*) (Table 1, Figure 2C). Regional plots are available in the online resources (Supplementary Figure S5-6).

### Functional interpretation and pleiotropic effect of genetic variants

We examined the potential causal variants within the identified SNPs (n=2,597) associated with coffee consumption, SNPs (n=14) associated with caffeine consumption, and SNPs (n=268) associated with other non-caffeine substances consumption, we found that the majority of these SNPs are located in intergenic and intronic areas (Supplementary Figure S7-9). Ninety-five SNPs (Supplementary Table 5) had likely deleterious impacts (CADD score >12.37) on gene functions ^18^. Six nonsynonymous among 95 SNPs located at gene exon region, including rs79217743 (*LMAN1L*), rs2231142 (*ABCG2*), rs35332062 (*MLXIPL*), rs6720 (*MDH2*), rs113534512 (*HCN2*), and rs1057868 (*POR*).

We also examined whether the identified SNPs are also eQTLs for nearby genes. The results were included in Supplementary Table 6. We found that 1941, 4 and 219 SNPs that are associated with the three phenotypes of GWAS 1, 2 and 3 are also significant eQTLs (FDR<0.05) for 180 genes in at least one tissue, respectively. For example, rs56113850-C associated with coffee consumption, which is also significantly associated with increased expression of both CYP1A6 and CYP1A7 (Supplementary Table 7). Novel risk alleles for coffee consumption, rs6655975-A, rs1571536-C, rs2667773-A, and rs3945-G, are associated with increased expression of *ADAMTSL4-AS1, GADD45G, LINGO1*, and *BRWD1*, respectively (Supplementary Table 7). Rs12898397-C, a missense variant with deleterious impact (CADD score=24.2) on *ULK3*, is associated with increased coffee consumption while a decreased expression of *ULK3* in multiple tissues (Supplementary Table 7). Rs11642015-T, associated with non-caffeine substances consumption, is associated with increased expression of FTO.

The gene enrichment analysis showed that these 180 GWAS SNP-associated genes were mainly involved in small molecule metabolic process, xenobiotic metabolic process, oxygenase p450 pathway, and generation of precursor metabolites and energy (Figure 3). Enrichment for tissue-specific expression of these genes showed that they were significantly overrepresented in heart and liver (FDR <0.1) (Figure 4).

**Figure 3.**
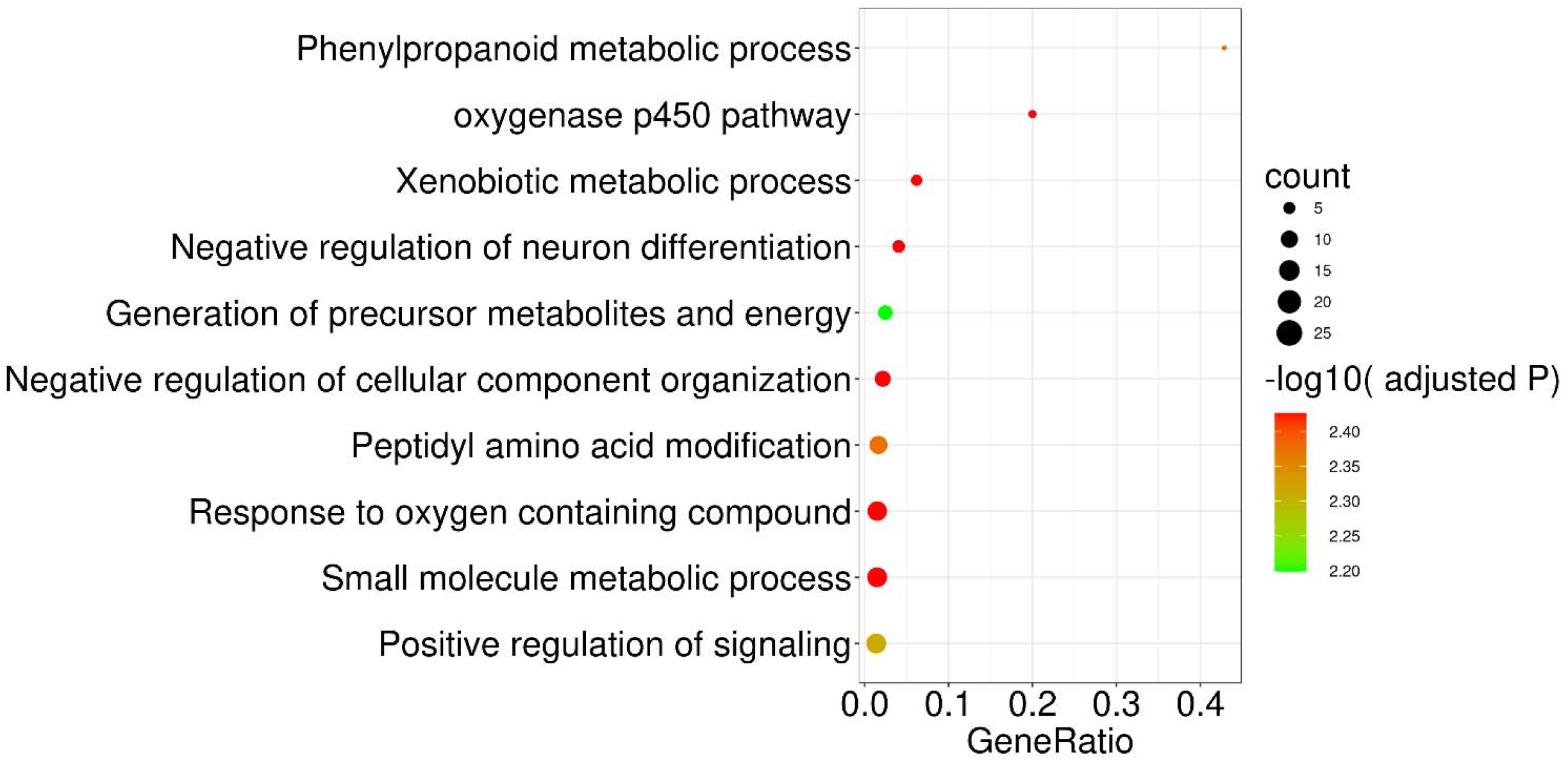
Pathway enrichment of the 180 genes associated with GWAS identified SNPs. The enrichment analysis was performed using GENE2FUNC in FUMA. The top 10 significantly enrichments (adjusted P <0.05) were available in the plot.

**Figure 4.**
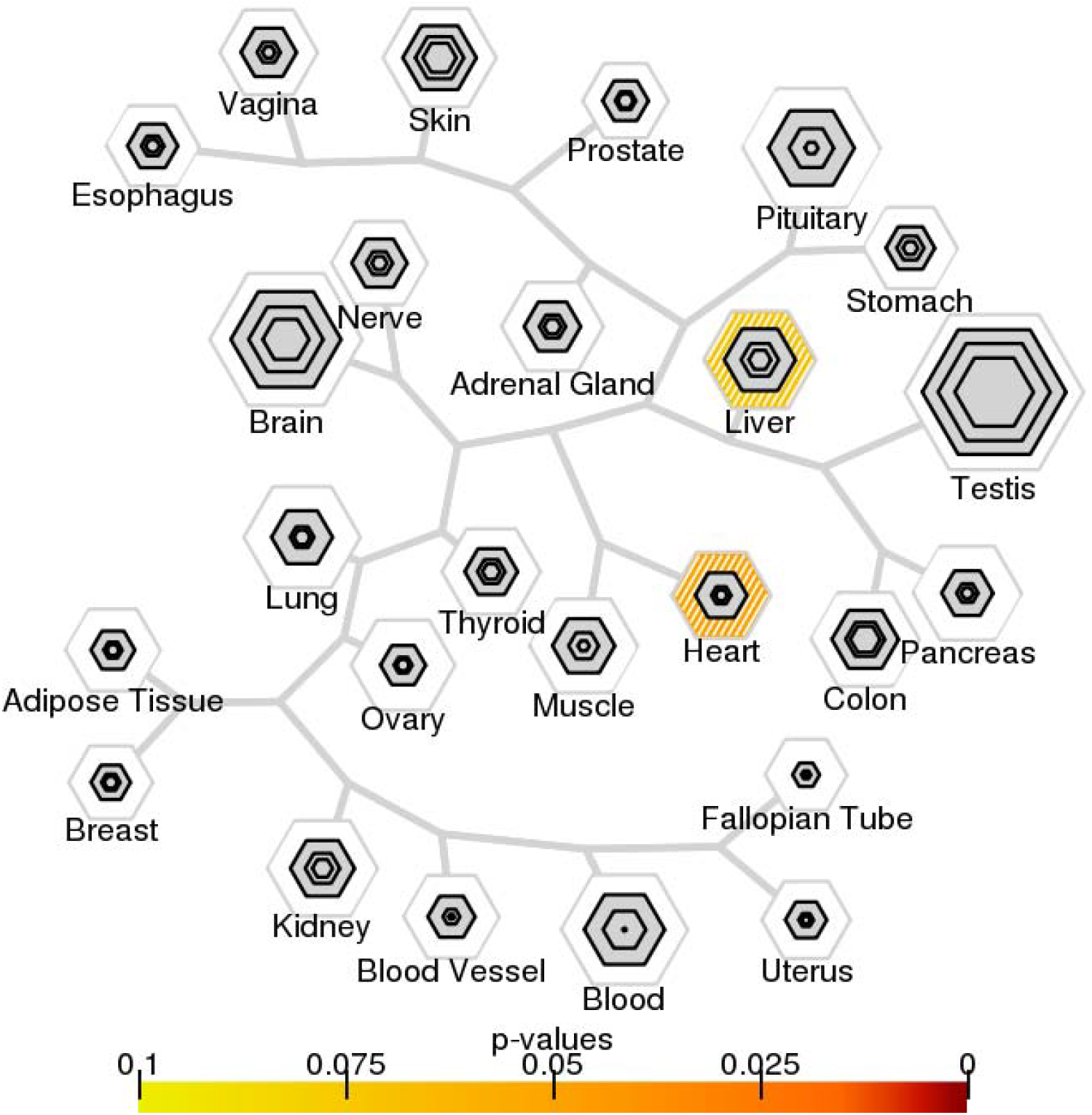
Expression enrichment analyses of the 180 genes associated with GWAS-identified SNPs. The tissue specific gene expression enrichment was analyzed using TSEA. Genes were significantly enriched in the liver and heart tissue (FDR <0.1).

Some of the lead SNPs alleles associated with the amount of coffee consumption were also associated with other traits in previously published GWA studies (Supplementary Tables 8). For instance, the alleles of rs2867110-G and rs476828-C were associated with increased BMI^27^; rs1260326-C, rs3792253-C and rs799166-G were associated with lowered triglycerides^28^; rs1467913-C,rs351237-G, rs12902040-T,rs9783698-G,and rs12914012-T were associated with lowered height^29; 30^. In addition, several alleles associated with higher coffee consumption were associated with decreased impedance of body (e.g. rs11078398-G), lowered creatinine in the urine (e.g. rs2472297-T), and increased age at menarche (e.g. rs3945-G)^30^. The lead SNP rs13107325-C associated with increased caffeine consumption was also associated with decreased body mass index^27^. The lead SNPs rs2067919-C and rs11642015-T associated with increased non-caffeine substances consumption were also associated with decreased alcohol intake frequency and increased risk of diabetes, respectively^30; 31^.

### Causal relationship between coffee consumption and the health consequences: TSMR

We used the 50 lead SNPs provided by our meta-analysis (Supplementary Table 4) as the IVs in our TSMR analysis. TSMR analyses involved 1,101 phenotypes in MR-Base as outcomes. For each outcome, only when the p-value of the IVs in the outcome is at least greater than 0.001, can it be used to infer the causality between coffee consumption and outcomes.

As shown in table 2, briefly we found significant causal relationships between coffee consumption and increased serum total cholesterol (id=933, beta=0.133 SD, p=1.27×10^−3^, FDR=1.86×10^−2^), serum total triglycerides (id=934, beta=0.154 SD, p=3.75×10^−4^, FDR=9.53×10^−3^), total cholesterol in LDL (id=895, beta=0.191 SD, p=4.49×10^−6^, FDR=9.34×10^−4^), apolipoprotein B (id=843, beta=0.233 SD, p=4.38×10^−8^, FDR=4.54×10^−5^), but decreased total cholesterol in HDL (id=864, beta=−0.128 SD, p=8.76×10^−4^, FDR=1.45×10^−2^). Furthermore, coffee consumption increased the risk of “less severe obesity”, including overweight (id=93, beta=0.124 log odds, p=8.87×10^−3^, FDR=7.75×10^−2^) and obesity class 1 (id=90, beta=0.195 log odds, p=4.45×10^−3^, FDR=4.43×10^−2^), but was not associated with obesity class 2 or 3. At the same time, waist circumference (id=61, beta=0.06 cm, p=9.21×10^−4^, FDR=1.45×10^−2^), waist-to-hip ratio (id=74, beta=0.09 SD, p=2.1×10^−4^, FDR>0.05), and body mass index (id=785, beta=−0.08 kg/m^2, p=7.19×10^−4^, FDR=1.29×10^−2^) were also increased with coffee consumption. Besides, coffee consumption was shown to increase the risk of type 2 diabetes (id=25, beta=1.15 log odds, p=3.18×10^−3^, FDR=3.45×10^−2^). In addition, the area under the curve (AUC) of insulin levels (id=760, AUCIns, beta=−0.250 mU*min/L, p=2.48×10^−2^, FDR>0.05), and corrected insulin response (id=761, CIR, beta=−0.206 SD, p= 3.21×10^−2^, FDR>0.05) during an OGTT were all decreased by coffee consumption. Other traits observed in our TSMR analysis can be found in Supplementary Table 9. Notably, coffee consumption showed differential risk on two types of ovarian cancer, with an increased risk for endometrioid ovarian cancer (id=1125, beta=0.349 log odds, p=1.84×10^−3^, FDR=2.37×10^−2^) but a decreased risk of low grade and low malignant serous ovarian cancer (id=1229, beta=−0.331 log odds, p=1.05×10^−2^, FDR>0.05).

**Table 2.** The causal associations between coffee consumption and human health outcomes based on the TSMR analysis.

| Outcome category | Outcome | Nsnp | BETA | SE | P | FDR | Heterogeneity | Pleiotropy |
| --- | --- | --- | --- | --- | --- | --- | --- | --- |
| lipid | Serum total cholesterol \|\| id:933 | 41 | 0.133 | 0.041 | $1.27 \times 10^{-3}$ | **$1.86 \times 10^{-2}$** | 0.261 | 0.599 |
| | Serum total triglycerides \|\| id:934 | 40 | 0.154 | 0.043 | $3.75 \times 10^{-4}$ | **$9.53 \times 10^{-3}$** | 0.155 | 0.628 |
| | Total cholesterol in LDL \|\| id:895 | 41 | 0.191 | 0.042 | $4.49 \times 10^{-6}$ | **$9.34 \times 10^{-4}$** | 0.233 | 0.939 |
| | Total cholesterol in HDL \|\| id:864 | 42 | -0.128 | 0.039 | $8.76 \times 10^{-4}$ | **$1.45 \times 10^{-2}$** | 0.642 | 0.746 |
| | Apolipoprotein B \|\| id:843 | 40 | 0.233 | 0.043 | $4.38 \times 10^{-8}$ | **$4.54 \times 10^{-5}$** | 0.247 | 0.635 |
| Anthropometric | Overweight \|\| id:93 | 27 | 0.124 | 0.047 | $8.87 \times 10^{-3}$ | $7.75 \times 10^{-2}$ | 0.118 | 0.889 |
| | Obesity class 1 \|\| id:90 | 27 | 0.195 | 0.069 | $4.45 \times 10^{-3}$ | **$4.43 \times 10^{-2}$** | 0.085 | 0.972 |
| | Waist circumference \|\| id:61 | 27 | 0.062 | 0.019 | $9.21 \times 10^{-4}$ | **$1.45 \times 10^{-2}$** | 0.278 | 0.569 |
| | Waist-to-hip ratio \|\| id:74 | 27 | 0.090 | 0.024 | $2.18 \times 10^{-4}$ | **$7.10 \times 10^{-3}$** | 0.192 | 0.861 |
| | Body mass index \|\| id:785 | 26 | 0.080 | 0.024 | $7.19 \times 10^{-4}$ | **$1.29 \times 10^{-2}$** | 0.154 | 0.919 |
| Diabetes | Type 2 diabetes \|\| id:25 | 2 | 1.154 | 0.391 | $3.18 \times 10^{-3}$ | **$3.45 \times 10^{-2}$** | 0.317 | NA |
| | AUCins \|\| id:760 | 28 | -0.249 | 0.111 | $2.48 \times 10^{-2}$ | $1.47 \times 10^{-1}$ | 0.309 | 0.023 |
| | Corrected insulin response \|\| id:761 | 27 | -0.206 | 0.096 | $3.21 \times 10^{-2}$ | $1.69 \times 10^{-1}$ | 0.650 | 0.207 |
| Cancer | Endometrioid ovarian cancer \|\| id:1125 | 43 | 0.349 | 0.112 | $1.84 \times 10^{-3}$ | **$2.37 \times 10^{-2}$** | 0.736 | 0.913 |
| | Serous ovarian cancer: low grade and low malignant potential \|\| id:1229 | 43 | -0.331 | 0.129 | $1.05 \times 10^{-2}$ | $8.66 \times 10^{-2}$ | 0.074 | 0.787 |

Footnotes: The outcomes causally associated with coffee consumption in our phenotype-wide TSMR analysis, which were categorized using the definitions in MR-Base, with a slight modification by merging the insulinemic outcomes into the “diabetes” category. The Outcome is the descriptive phenotype with a unique id assigned by MR-Base; Nsnp: the number of IVs used in TSMR; BETA: the causal effect of coffee consumption on outcome; SE: the standard error; P: the p-value of TSMR analysis; FDR: false discovery rate; Heterogeneity: the p-value of Cochran’s Q value; Pleiotropy: the p-value of MR-Egger intercept.

To reveal the diseases categories or traits which are mostly affected by coffee consumption, we conducted an enrichment analysis of the significant causal effects. The results showed that the health outcomes causally driven by the coffee consumption were significantly enriched in several MR-base categories, i.e. blood lipids, fatty acids or amino acids in plasma/serum, and anthropometric measurements (Figure 5).

**Figure 5.**
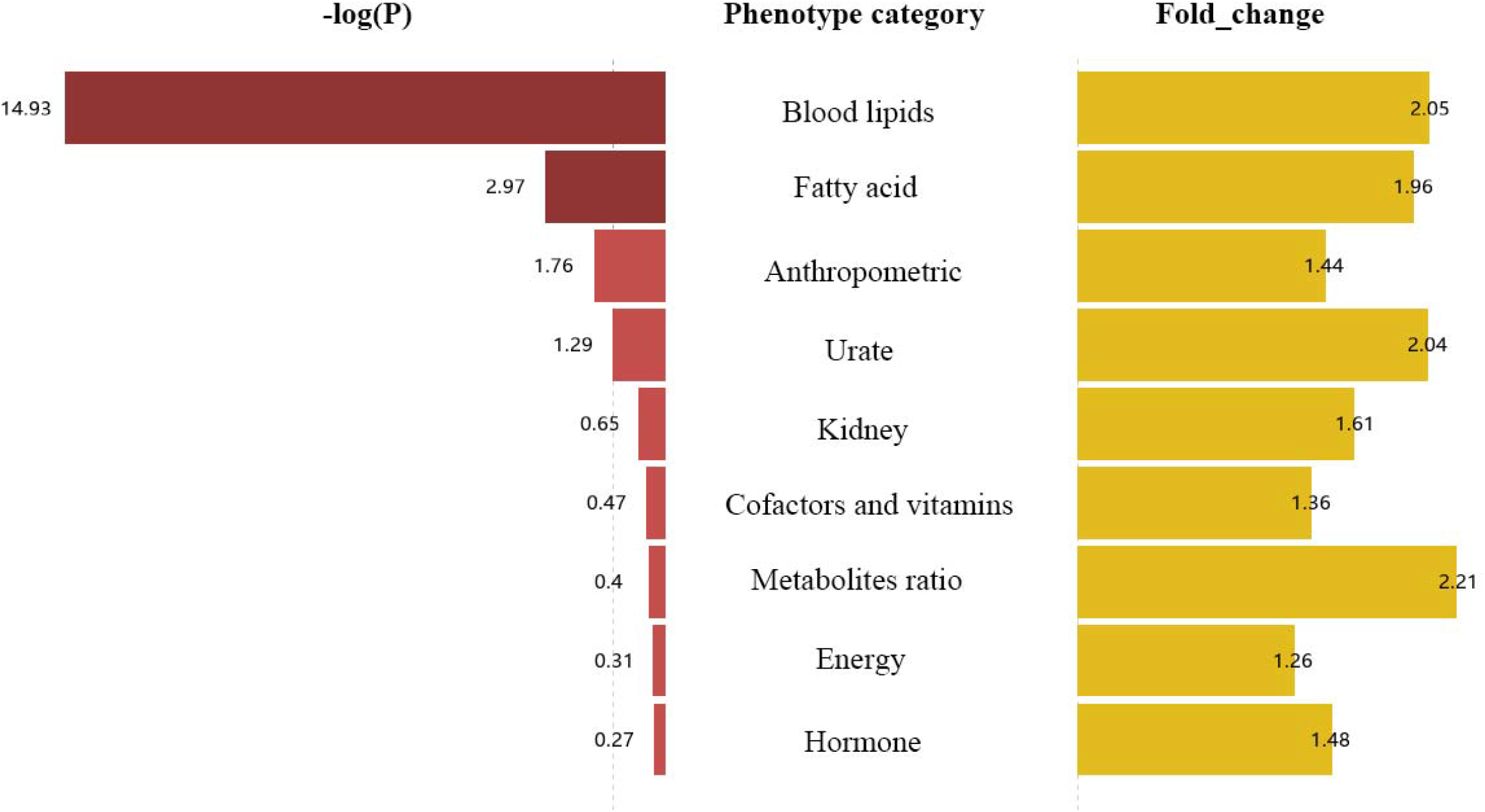
The enrichment analysis of the outcomes causally associated with the genetically driven coffee consumption among the phenotypic categories of various traits defined in MR-Base. For each category, the −log_10_ of the enrichment p value was indicated on the left, while the enriched fold change was indicated on the right. The phenotype categories with FDR ≤0.05 are highlighted in dark red.

### Validation of the associations between coffee consumption and health outcomes: OSMR

We used the IVs actually used in each outcome in the TSMR to construct the PRS score separately and inferred the relationship with the corresponding outcomes.

As shown in table 3, OSMR analyses were performed to further validate the significant findings in TSMR. In general, our results showed a consistent association between coffee consumption and similar metabolic traits as noted above. Briefly, the coffee-PRS was positively associated with cholesterol (p=1.65×10^−4^), LDL (p=7.76×10^−6^), apolipoprotein B (p=1.24×10^−5^), but negatively associated with HDL (6.56×10^−3^). Furthermore, coffee-PRS was positively associated with waist circumference (p=4.08×10^−10^), hip circumference (p=7.56×10^−9^), body mass index (p=3.18×10^−15^) and weight (p=2.66×10^−8^). Coffee-PRS was also positively associated with glycated haemoglobin (HbA1c, p= 3.28×10^−6^) and diabetes (p=2.00×10^−4^). Lastly, coffee-PRS also showed a trend of negative association with ovarian cancer (p=4.80×10^−2^).

**Table 3.** The causal associations between coffee consumption and human health in UKBB based on OSMR analyses.

| Outcome category | Outcome | N (case \| control) | BETA | SE | P |
| --- | --- | --- | --- | --- | --- |
| Age | Age | 193674 | 7.407 | 6.264 | $2.37 \times 10^{-1}$ |
| Sex | Sex | 193674 | 0.053 | 0.393 | $8.92 \times 10^{-1}$ |
| Lipids | Cholesterol | 184858 | 3.333 | 0.885 | **$1.65 \times 10^{-4}$** |
|  | LDL | 184540 | 3.013 | 0.674 | **$7.76 \times 10^{-6}$** |
|  | Apolipoprotein B | 183961 | 0.788 | 0.180 | **$1.24 \times 10^{-5}$** |
|  | HDL | 169307 | -0.862 | 0.317 | **$6.56 \times 10^{-3}$** |
| | Apolipoprotein A | 168330 | -0.412 | 0.220 | $6.08 \times 10^{-2}$ |
| Anthropometric | Waist circumference | 193635 | 50.085 | 8.012 | **$4.08 \times 10^{-10}$** |
|  | Hip circumference | 193634 | 31.293 | 5.416 | **$7.56 \times 10^{-9}$** |
|  | Body mass index | 193674 | 21.551 | 2.733 | **$3.18 \times 10^{-15}$** |
|  | Weight | 193674 | 52.653 | 9.465 | **$2.66 \times 10^{-8}$** |
| Diabetes | Glucose | 169192 | 0.411 | 0.603 | $4.95 \times 10^{-1}$ |
|  | Glycated haemoglobin (HbA1c) | 184763 | 19.021 | 4.088 | **$3.28 \times 10^{-6}$** |
|  | Diabetes | 9367 \| 183926 | 0.334 | 0.090 | **$2.00 \times 10^{-4}$** |
| Cancer | Ovarian cancer | 885 \| 101366 | -0.195 | 0.098 | **$4.80 \times 10^{-2}$** |

Footnotes: Outcome categories based on the definitions in MR-Base. Similarly, we also classified glycemic outcomes into the “diabetes” category. N, the number of participants used in OSMR; BETA, the regression coefficient of coffee consumption PRS; SE, the standard error of BETA; P: the p-value of OSMR analysis.

### Causal relationship between the consumption of caffeine and non-caffeine components in coffee and metabolic traits

We set out to examine which components in coffee may lead to the potential detrimental effects of coffee consumption on metabolic perturbations. We conducted TSMR using 38 SNPs (Supplementary Table 10) from GWAS 2 and 83 SNPs (Supplementary Table 11) from GWAS 3 with p<10^−5^ as a liberal cut-off for selecting the IVs, to examine the effects of caffeine or other non-caffeine components in coffee on coffee-consumption associated metabolic traits identified in the aforementioned TSMR analysis. We found that caffeine exposure was negatively associated with concentration of chylomicrons and largest VLDL particles (id=958, beta= −1.128 SD, p=2.76×10^−2^), and concentration of medium VLDL particles (id=913, beta=−0.976 SD, p=3.18×10^−2^). While the intake of other components in coffee increased the total cholesterol in LDL (id=895, beta=0.320, p=3.50×10^−2^) (Supplementary Table 12).

## Discussion

We performed the largest-to-date GWA and meta-analysis on coffee consumption. We further for the first time conducted GWA studies on caffeine intake and decaffeinated coffee intake. Our analyses identified novel loci associated with each of the three phenotypes, which provide new insights into the genetic basis underlying the coffee consumption behavior among human populations. Moreover, by leveraging these genetic findings, we performed large-scale MR analyses to assess the causal relationship between different coffee intake behavior and the health outcomes. Our study indicated that, unlike what have found in many observational studies^1-4; 32^, coffee consumption may causally lead to increased risks for metabolic diseases, and the more coffee consumed, the worse the outcomes. This finding is consistent with several studies based on RCTs^5-9^, having important implications for public health.

In our GWA studies, we confirmed all 10 known^11^ and newly identified 9 genetic loci associated with coffee consumption among a large, combined European population. A recent GWAS also using UKBB samples identified fewer loci associated with coffee consumption^25^. The discrepancy between this GWAS^25^ and our coffee consumption GWAS 1 may be attributed to the sample selection and inclusion of different covariates. The former study included both coffee and non-coffee drinkers in UKBB European population^25^, while our GWAS 1 was restricted to the coffee drinkers of European-ancestry.

Furthermore, statistical models of the former adjusted for age, sex, BMI and top 20 principal components^25^, while our statistical models additionally adjusted for coffee type and smoking. These settings allow us to combine the CCGC GWAS study to perform the meta-analysis. In addition, we also identify 2 and 5 novel loci significantly associated with caffeine and non-caffeine coffee components. The enrichment analysis of the genes whose transcription is associated with these SNPs revealed pathways related to small molecule metabolic process, xenobiotic metabolic process, oxygenase p450 pathway, etc., highlighted that genetic variants altering the metabolism of caffeine and related active xenobiotic compounds in coffee are likely the major determinants for coffee consumption behaviors.

This is consistent with the previous identification^11^. Meanwhile, these GWAS SNP-regulated genes are enriched in the liver and heart tissue, with also a significant enrichment related to the generation of precursor metabolites and energy (Figure 4), suggesting a potentially overlap between the coffee consumption and energy metabolism. In addition, our genome-wide meta-analysis of coffee consumption identified candidate SNPs that may have deleterious impacts (CADD score >12.37) on gene functions. In particular, rs12898397-C (CADD score =24.2), a missense variant with deleterious impact on *ULK3* (Unc-51 like kinase 3), was associated with increased coffee consumption but decreased expression of *ULK3*. ULK3 is a serine/threonine protein kinase that acts as a regulator of sonic hedgehog (SHH) signaling and autophagy. *ULK3* low expression may induce the dysregulation of autophagy, which participates in controlling the metabolic functions of liver via multiple ways^33^.

We performed GWA studies on caffeine consumption and non-caffeine substances consumption, which have never been investigated at the genome-wide level. We found that rs112764911 (*SORCS2*) and rs13107325 (*SLC39A8*) were associated with caffeine consumption, and the latter may be also associated with coffee consumption (p=2.48×10^−4^). Interestingly both genes were identified among a number of GWA studies to be associated with multiple phenotypes especially neuropsychiatric diseases and traits, e.g. SNPs in *SORCS2* are highly expressed in the brain tissue, and previously was associated with attention function in attention deficit hyperactive disorder^34^, general risk tolerance and risk behavior^35^, alcohol withdrawal symptom^36^, depressive and manic episodes in bipolar disorder^37^, etc.; while SNPs in *SLC39A8* were also associated with schizophrenia^38^, bipolar disorder^39^, and intelligence^40^, etc., indicating the impact of caffeine on central nerve system and the potential connection between caffeine intake and neuropsychiatric reactions. In addition, we identified 5 loci (*LINC02060, AHR, PRR4, CYP1A1*, and *FTO)* associated with drinking of decaffeinated coffee. Two of them (*AHR* and *CYP1A1*) are also associated with coffee consumption in GWAS. AHR is known to be activated by many xenobiotic compounds, e.g. polycyclic aromatic hydrocarbons (PAHs) in coffee^41^. AHR response elements reside in the bidirectional promoter region located at chromosome 15q24, which associated with transcriptional activation of both *CYP1A1* and *CYP1A2*^42-44^. While CYP1A1 plays an important role in metabolizing and the detoxification of PAHs, CYP1A2 directly metabolizes caffeine^45^, which may explain the overlapping identification between the two GWA studies. In addition, the *FTO* gene is known to be associated with BMI and involved in energy metabolism, further indicated the close connection between coffee intake and energy intake/homeostasis. Our study warrants continued investigations for the detailed mechanism underlying how these genes determining the caffeine or decaffeinated coffee intake.

In order to investigate the potential impact of regular consumption of coffee, caffeine or non-caffeine coffee constituents on human health, we performed MR analyses and found that coffee consumption may causally lead to altered risks for multiple clinical outcomes which are enriched in metabolic perturbations, especially increased risks for dyslipidemia, obesity, and diabetes. Meanwhile, non-caffeine substances consumption increased the risks for high blood lipids, while caffeine consumption decreased the risks for high blood lipids. The impact of coffee on human health has been long speculated to be attributed to the various bioactive components contained in coffee, such as caffeine, chlorogenic acid, diterpenoids, PAHs, etc. In general, due to the bitter taste of coffee, coffee consumption is more likely to be associated with increased intake of sugary, thereby increase the risk of diabetes and obesity^46; 47^. Meanwhile, after consumers switch from caffeinated coffee to decaffeinated coffee, LDL cholesterol and apolipoprotein B increase, suggesting that other coffee components other than caffeine may be responsible for the high blood lipids^48^. Consistent with our findings, caffeine has been demonstrated to have a beneficial impact on lipid metabolism, which reduces intrahepatic lipid content and stimulates β-oxidation in hepatic cells and liver via regulating the autophagy-lysosomal pathway signaling^49^. While, diterpenoids contained in coffee may be an important factor leading to the increase of blood lipid level, and its impact on increasing blood lipid level may be related to its impact on the activity of serum lipid transporters^50^. A study tested the effect of cafestol, a diterpenoid in coffee, by giving to 10 healthy male volunteers for 28 days. Relative to baseline values, cafestol raised the activity of cholesterylester transfer protein by 18 +/−12% and of phospholipid transfer protein by 21 +/−14% (both P < 0.001), which may be associated with elevated serum VLDL and LDL cholesterol^50^.

Furthermore, filtered coffee containing a relative lower amount of diterpenoids does not increase blood lipid levels^51; 52^. In addition, an observational study showed that C-peptide, a marker of insulin secretion, decreased with every additional cup of decaffeinated coffee (0.063 ng/ml; P = 0.0003), which indicated the potential function of non-caffeine substances for diabetic risk^53^. Taken together, our data suggest that the detrimental effects of coffee consumption on health outcomes may be due to the intake of non-caffeine components. While even though caffeine may exert some beneficial impact on lipids homeostasis, this impact may not be sufficient to compensate the deleterious effects of the non-caffeine components in the coffee. The detailed mechanism underlying the association between coffee consumption and metabolic perturbations remains to be further investigated.

It is noteworthy that our MR analyses also suggested a potential causal relationship between coffee consumption and ovarian cancer, with an opposing risk for different subtypes of the disease. While the detailed mechanism underlying this association remains unclear. Also, caffeine has also been demonstrated to decrease estrogen but increase progesterone, which has been weakly associated with ovarian cancer risk^54; 55^. More studies are needed to further clarify this relationship.

## Limitations

The current study was conducted in the participants of European ancestry. Whether our findings can be generalized to other ethnic groups remains to be validated in future studies. Moreover, although the study has identified 19 genomic loci (9 of which are novel loci) associated with coffee consumption, they only explain a small proportion of the phenotype variance, indicating that our MR results were driven by a limited proportion of genetic susceptibility. Our conclusion may not represent the full image of the health impact of coffee consumption. Meanwhile, the findings of GWAS 2 and 3 studies need further independent validations as well.

## Conclusions

This study identified novel genetic loci associated with multiple coffee consumption behaviors and provided evidence that coffee consumption increases the risk for metabolic diseases. This could have significant implications for global public health given the increasing burden of metabolic diseases.

## Data Availability

The study was conducted using the UK Biobank data resources under application number 53536.The coffee consumption GWAS for genome-wide meta-analysis was provided by the Coffee and Caffeine Genetics Consortium.A large number of phenotype previously studied among various GWAS were provided by MR-Base.

https://digitalhub.northwestern.edu/

https://www.mrbase.org

## Acknowledgements

This work was supported by the “Changbai Mountain Scholar” Distinguished Professor Awarding Program of the Department of Education of Jilin Province, China. This work was supported in part by the Start-Up Fund (W.L) of the Wayne State University.

## Author contributions

J.L, W.L and P.C conceived the study. P.C contributed to the acquisition of UKBB data. J.L, M.Z and L.C analyzed the data. J.L, W.L, J.W and P.C interpreted the results. J.L, T.C and W.L wrote the first draft of the manuscript. All authors revised the manuscript and approved the submission.

## Notes

### Competing Interest Statement

The authors have declared no competing interest.

### Author Declarations

This study was conducted using the UKB resources under the application 53536. The UK Biobank was approved by the North West Multi-center Research Ethics Committee, the National Information Governance Board for Health and Social Care in England and Wales, and the Community Health Index Advisory Group in Scotland. Written informed consent was obtained from all participants at recruitment.

### Summary of Updates

Figure 1 revised.

