## Supplementary figures for "Habitual Coffee Consumption Increases Risks for Metabolic Diseases: Genome-wide Association Studies and a Phenotype-wide Two Sample Mendelian Randomization Analysis": Supplementary figures.docx

**Supplementary Figure 1. QQ plot for** **genome-wide meta-analysis of coffee consumption among 375388 coffee consumers.** The QQ plot displays the expected –log10 (p-values) on the x-axis and the observed –log10 (p-values) on the y-axis.

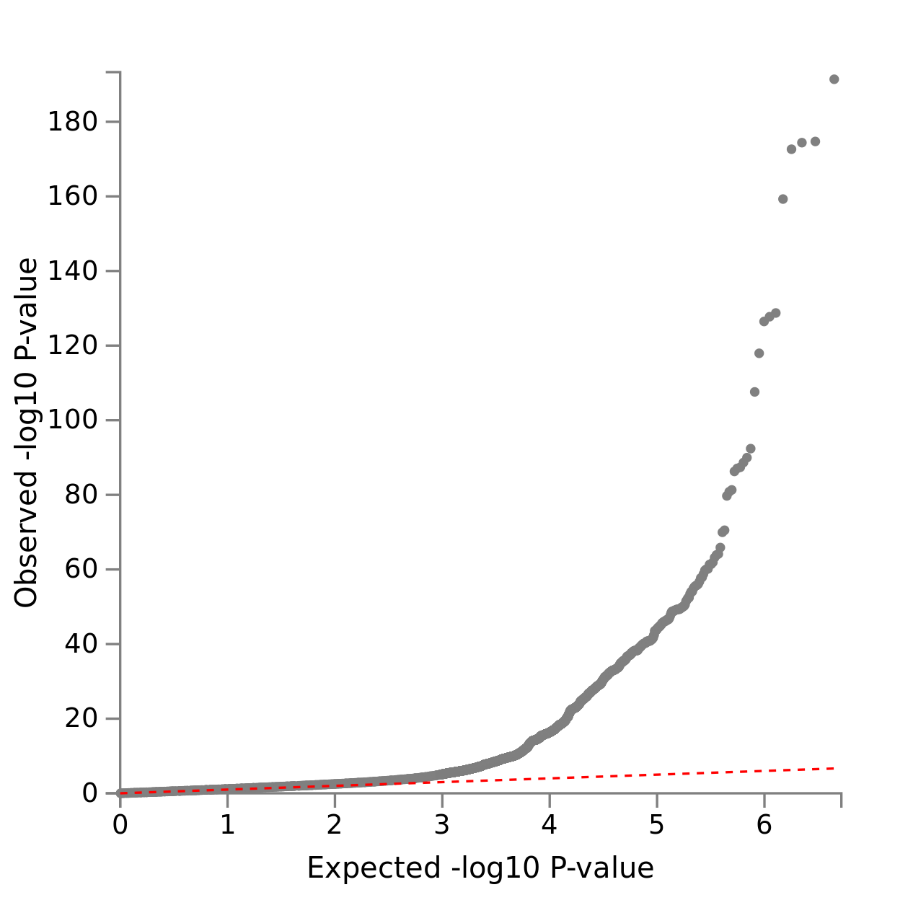

**Supplementary Figure 2. Regional plot for the 19 significant loci of coffee consumption.** Each plot represents a SNP, which are color-coded based on the highest r2 to one of the leading SNPs. SNPs with r2 < 0.05 are coloured in grey. The red coloured gene were mapped by FUMA using positional mapping.

a. locus 1

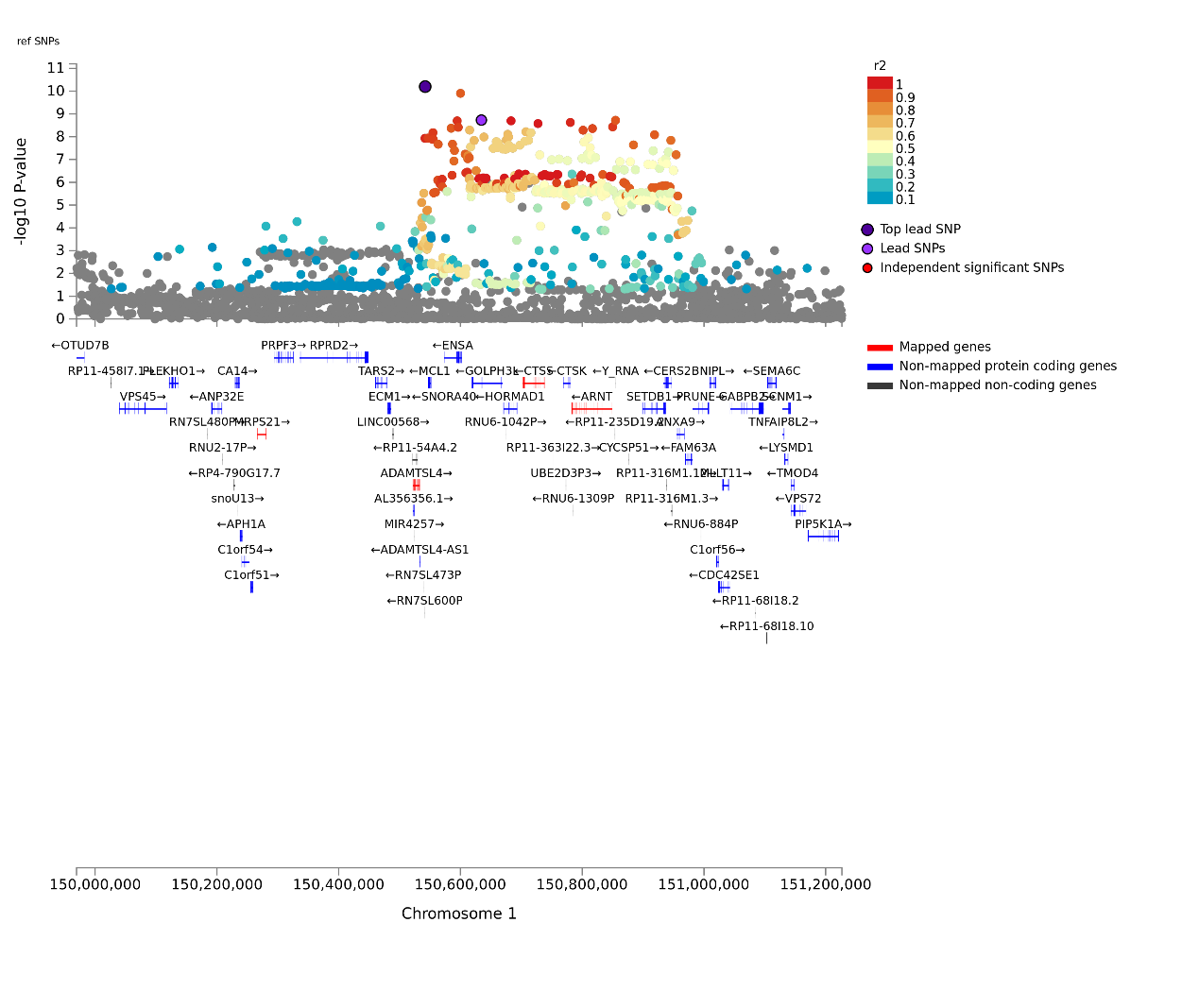
b. locus 2

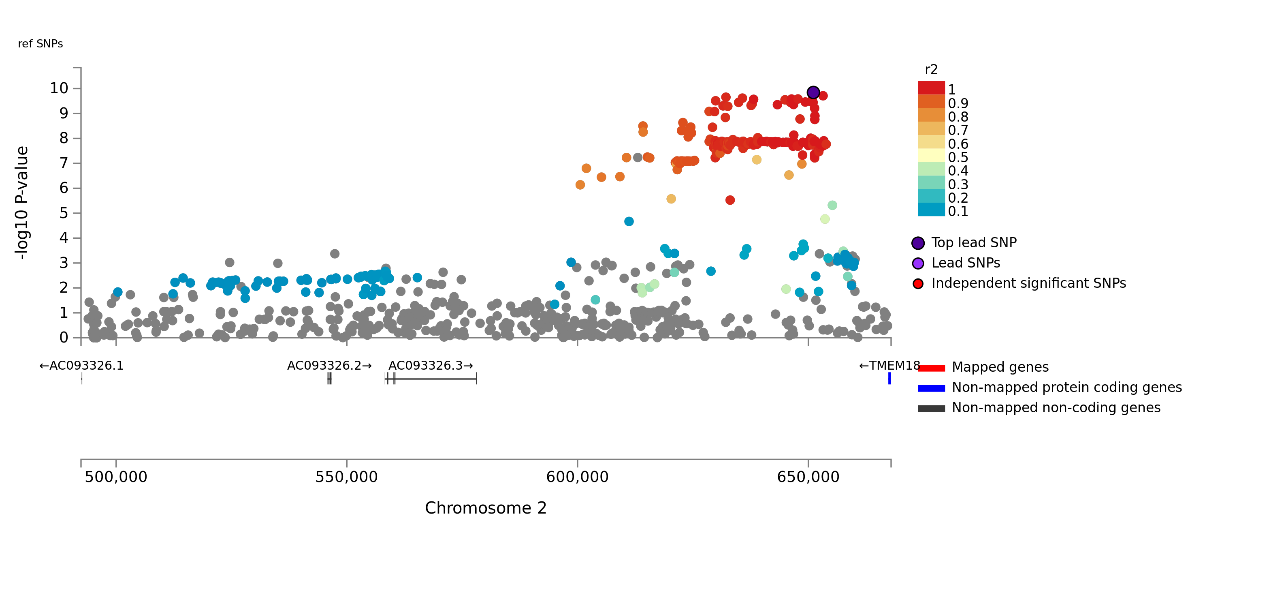

c. locus 3

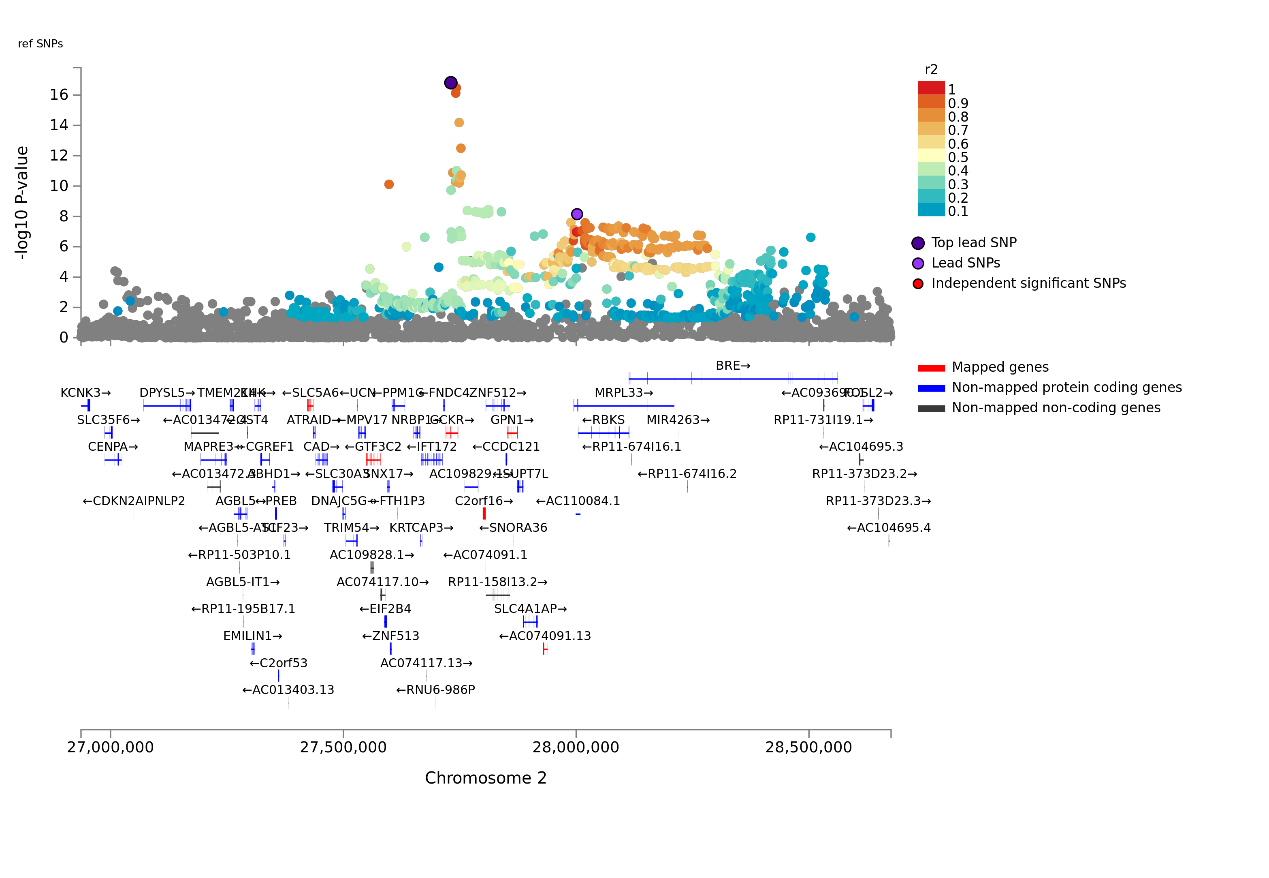

d. locus 4

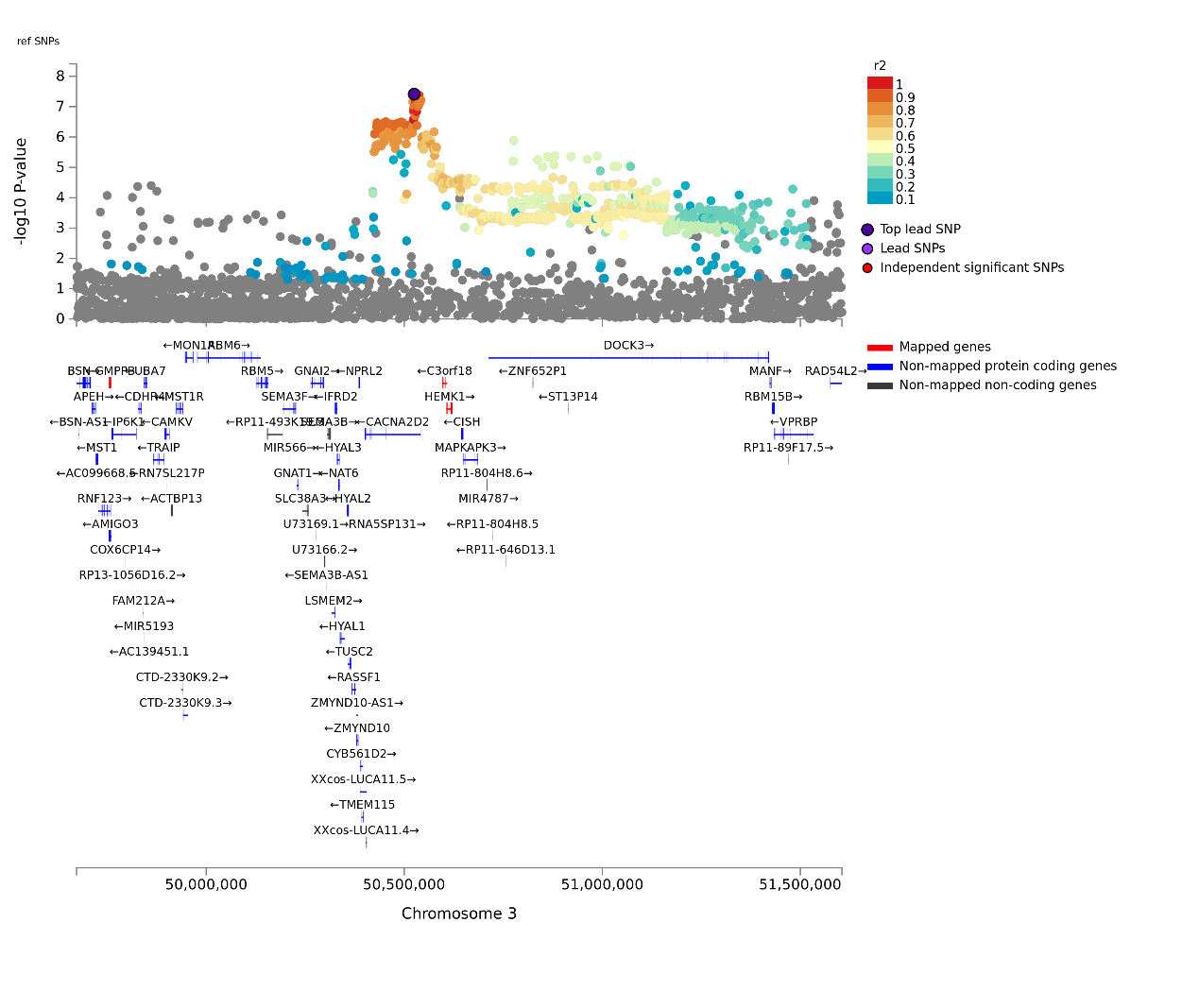
e. locus 5

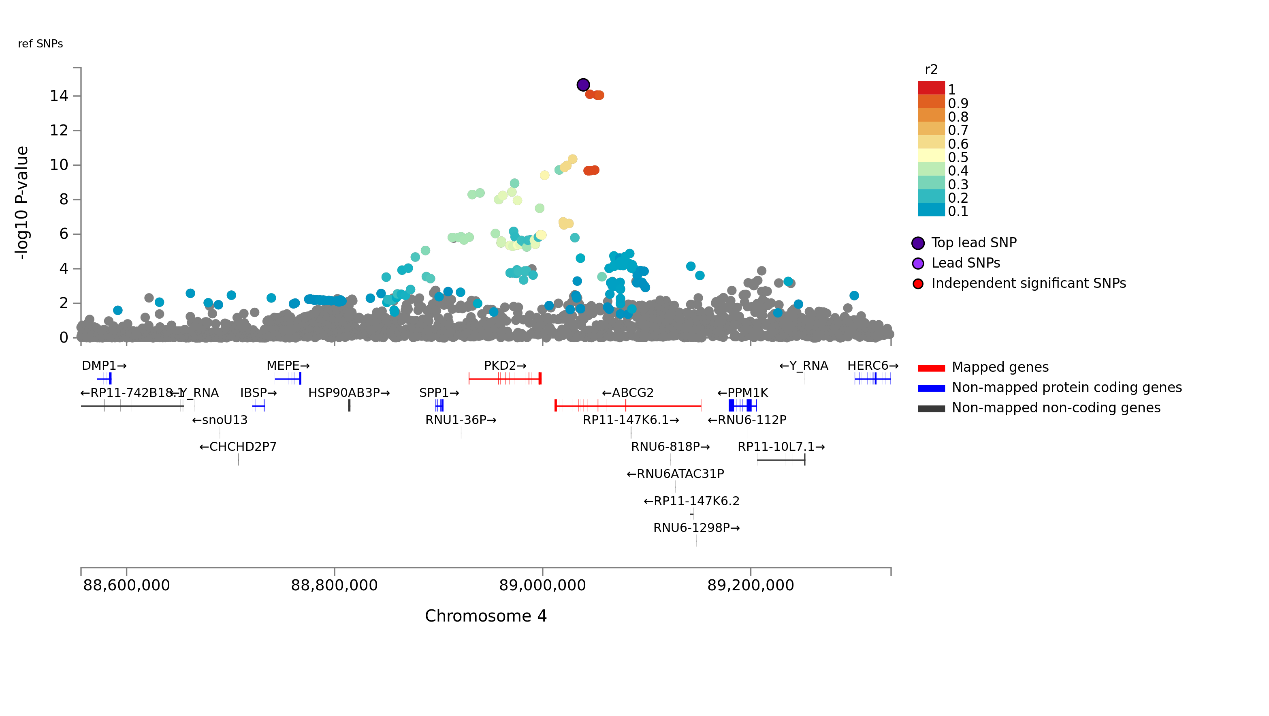
f. locus 6

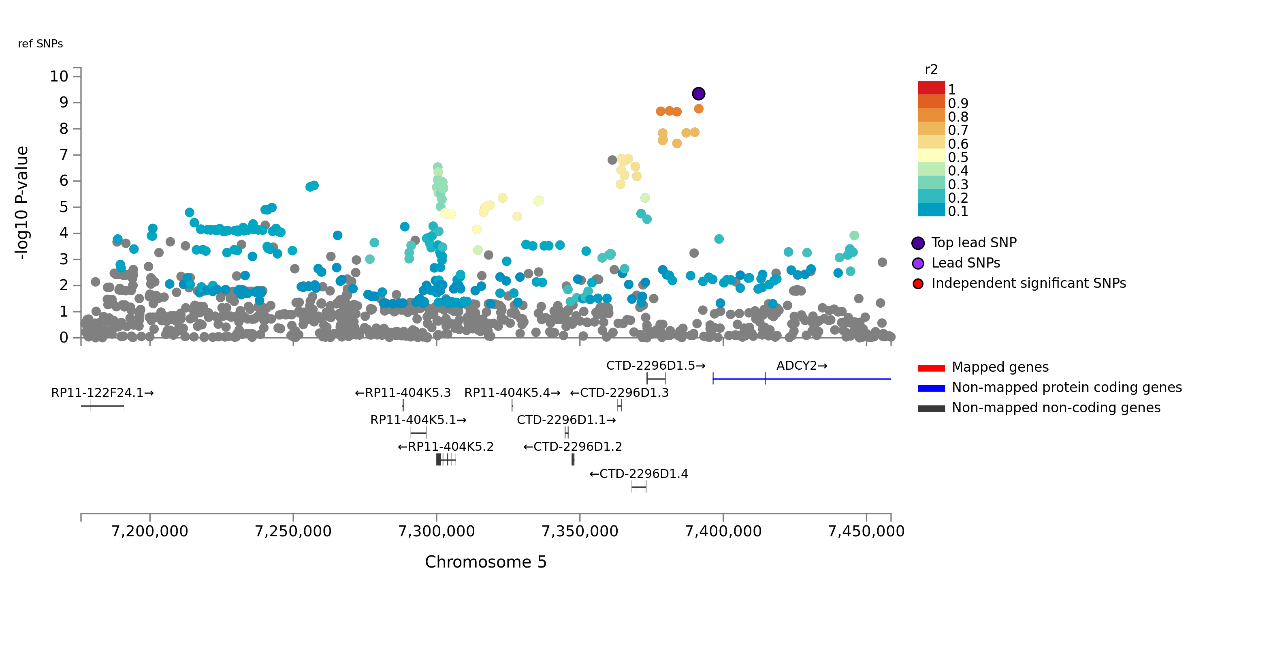

g. locus 7

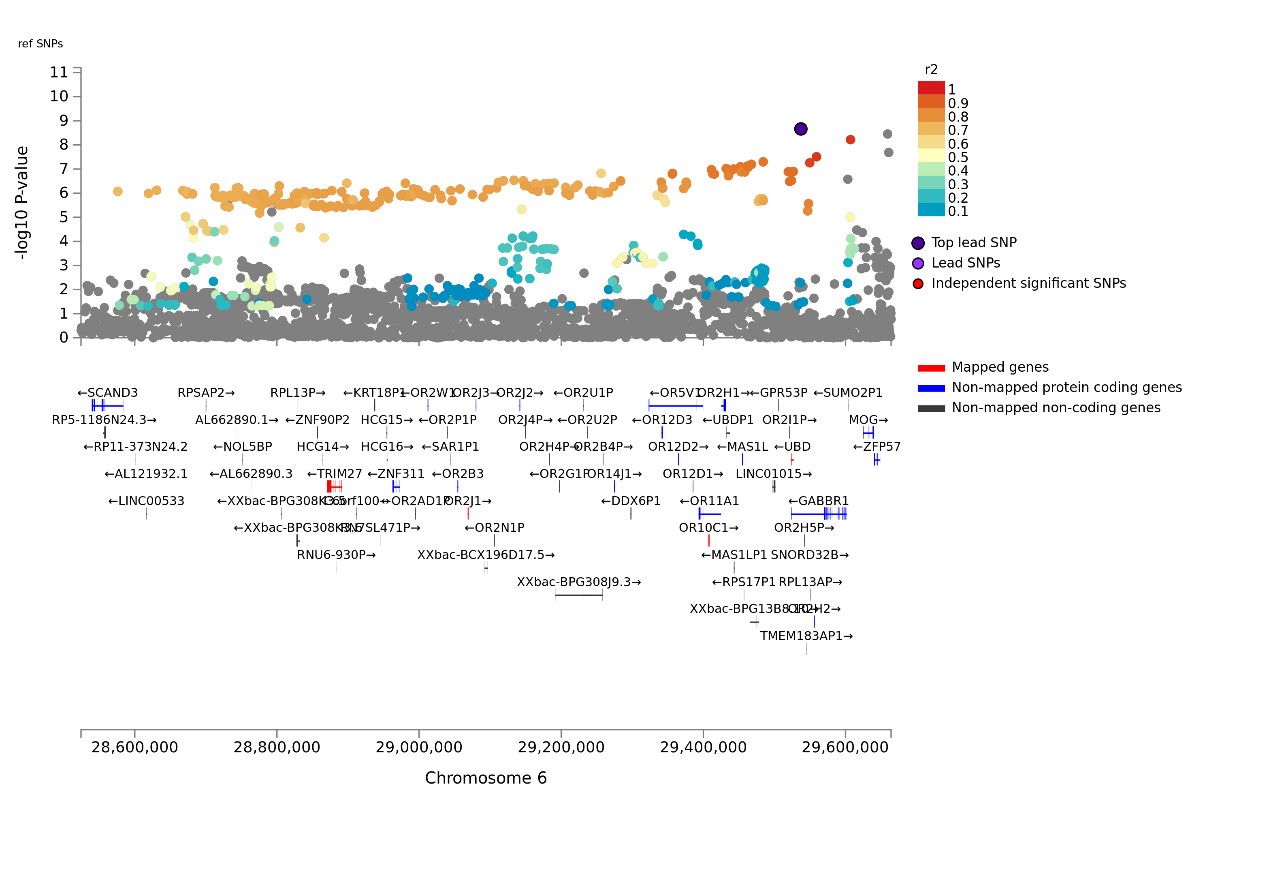

h. locus 8

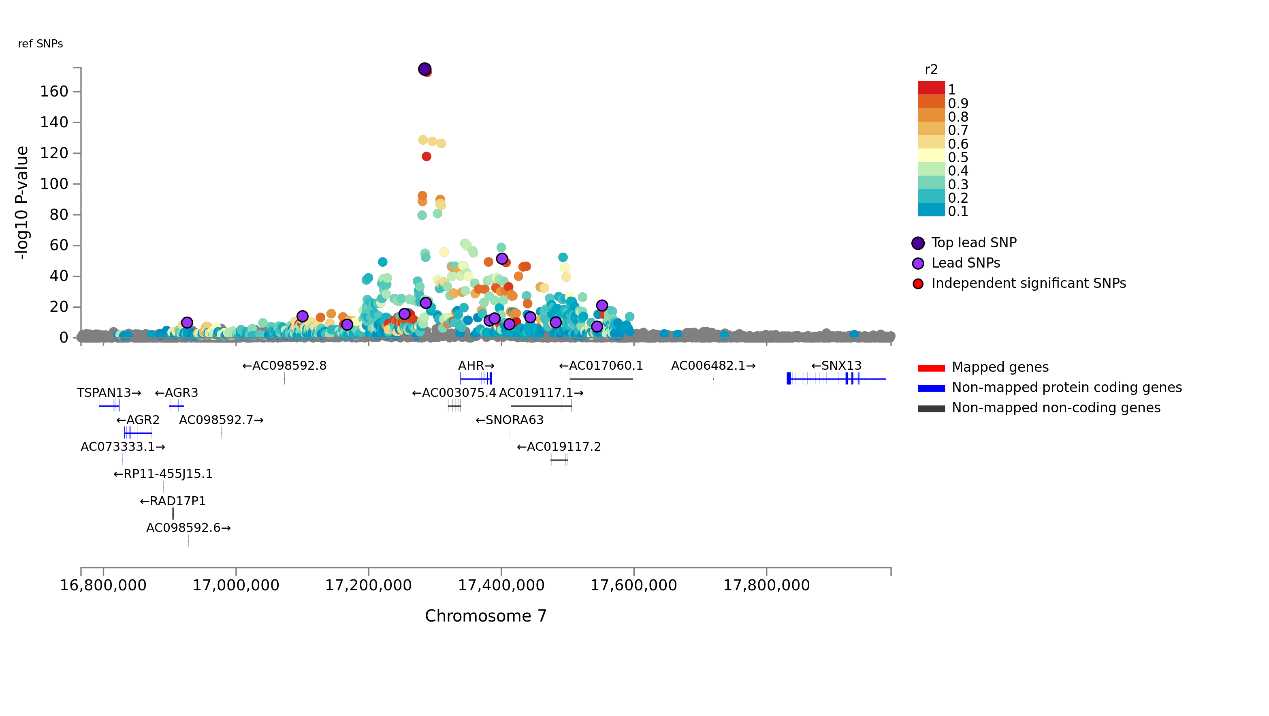

i. locus 9

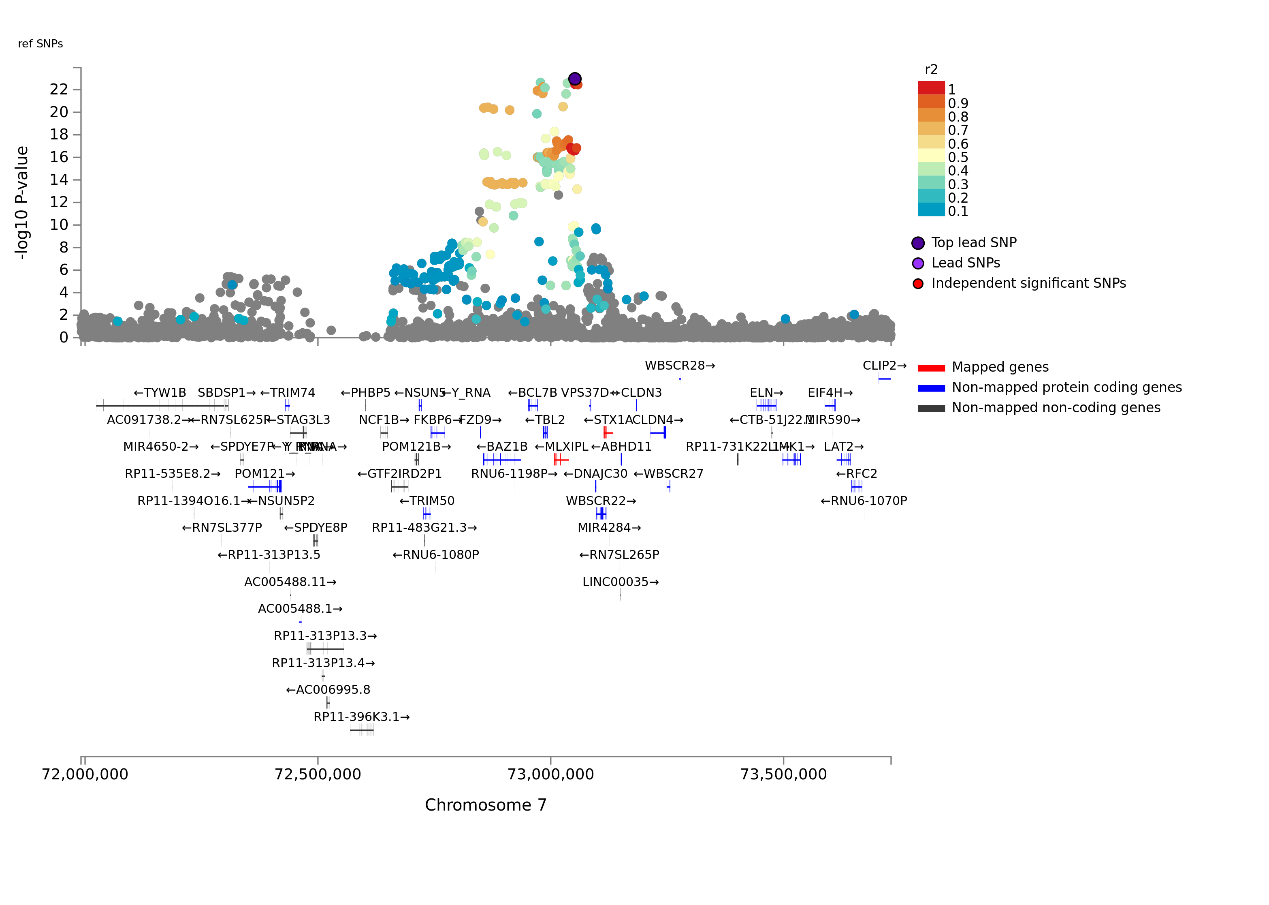
j. locus 10

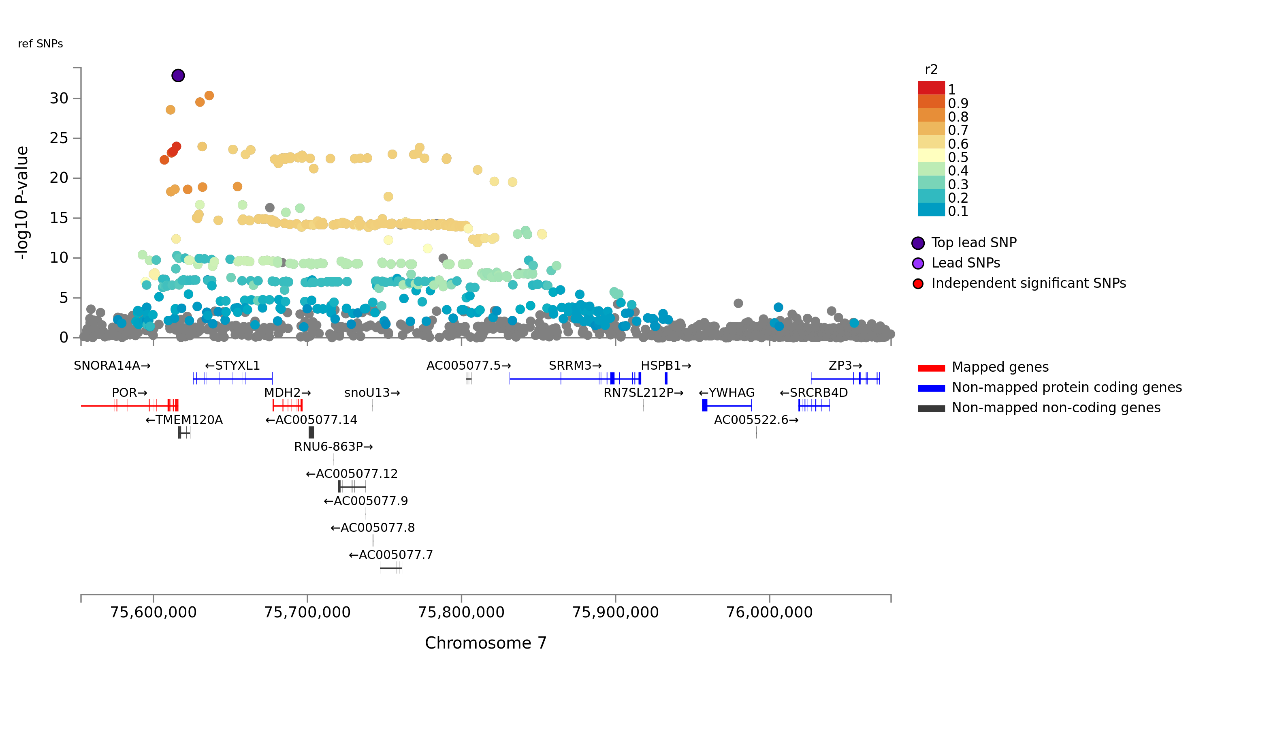

k. locus 11

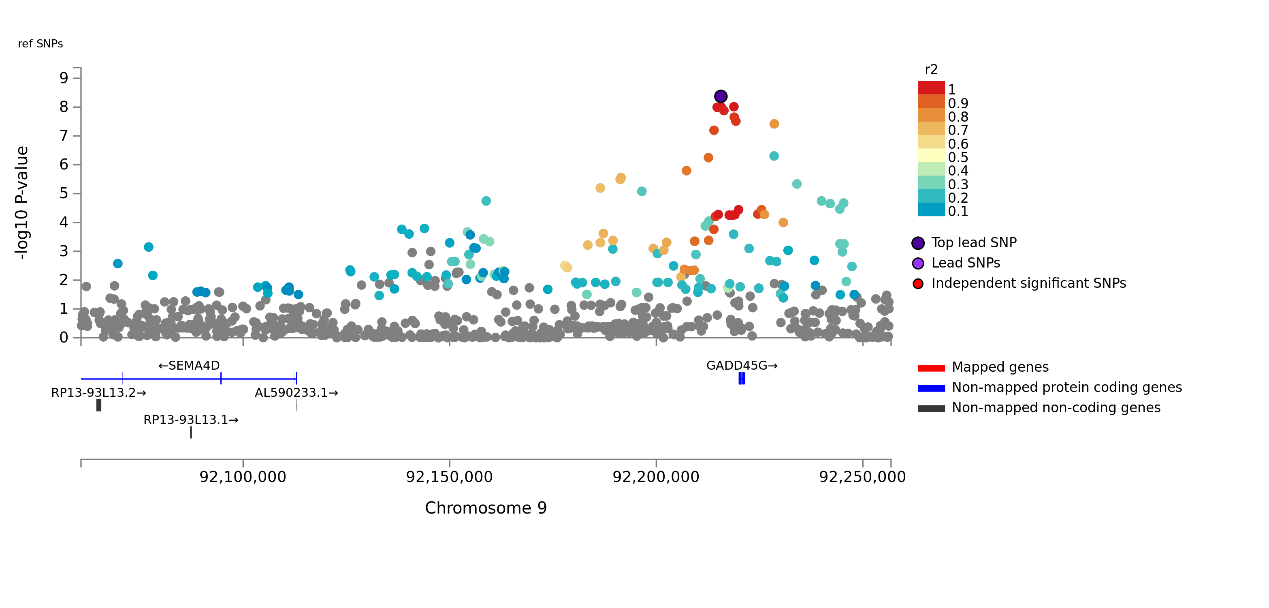

l. locus 12

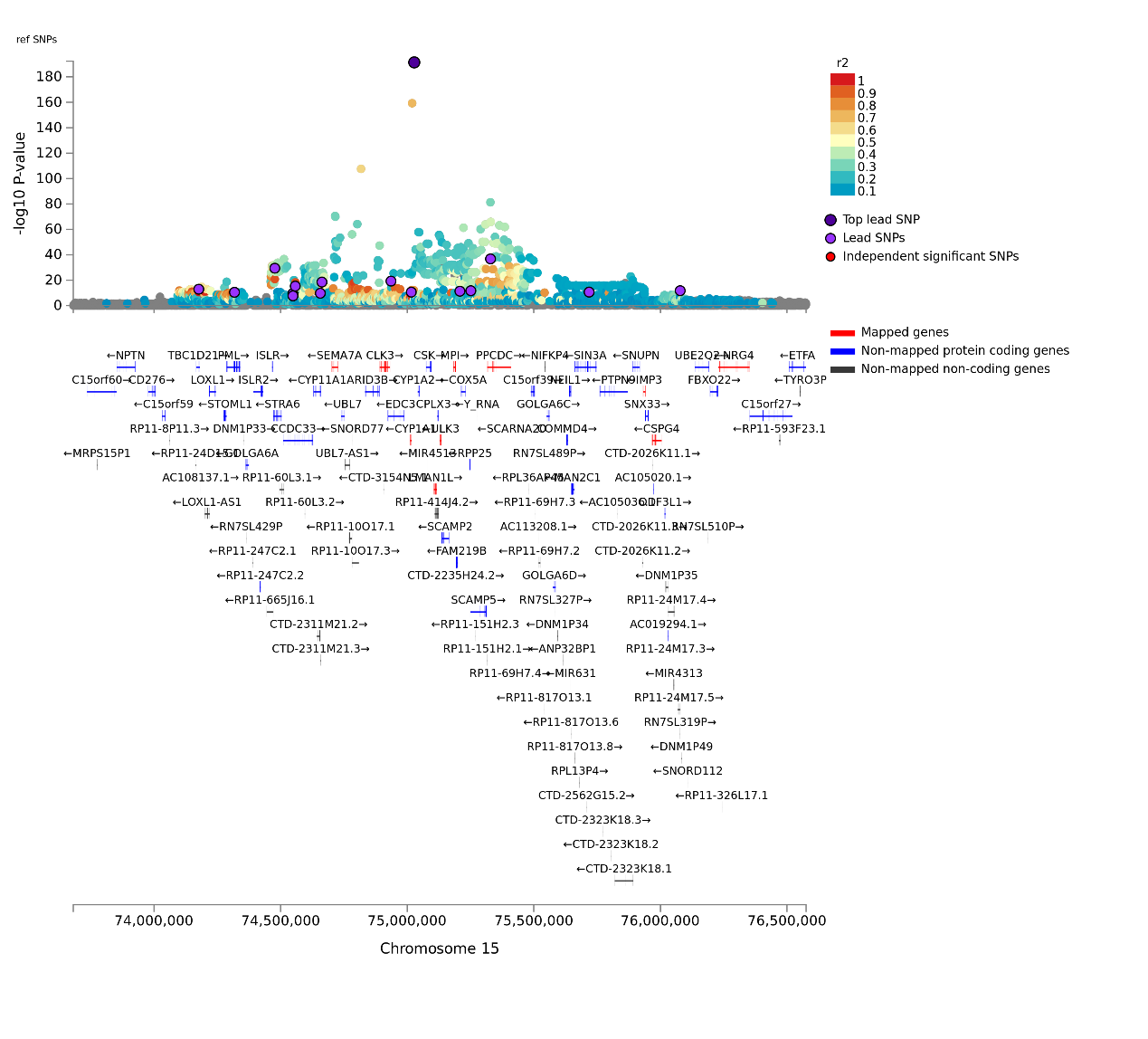

m. locu13

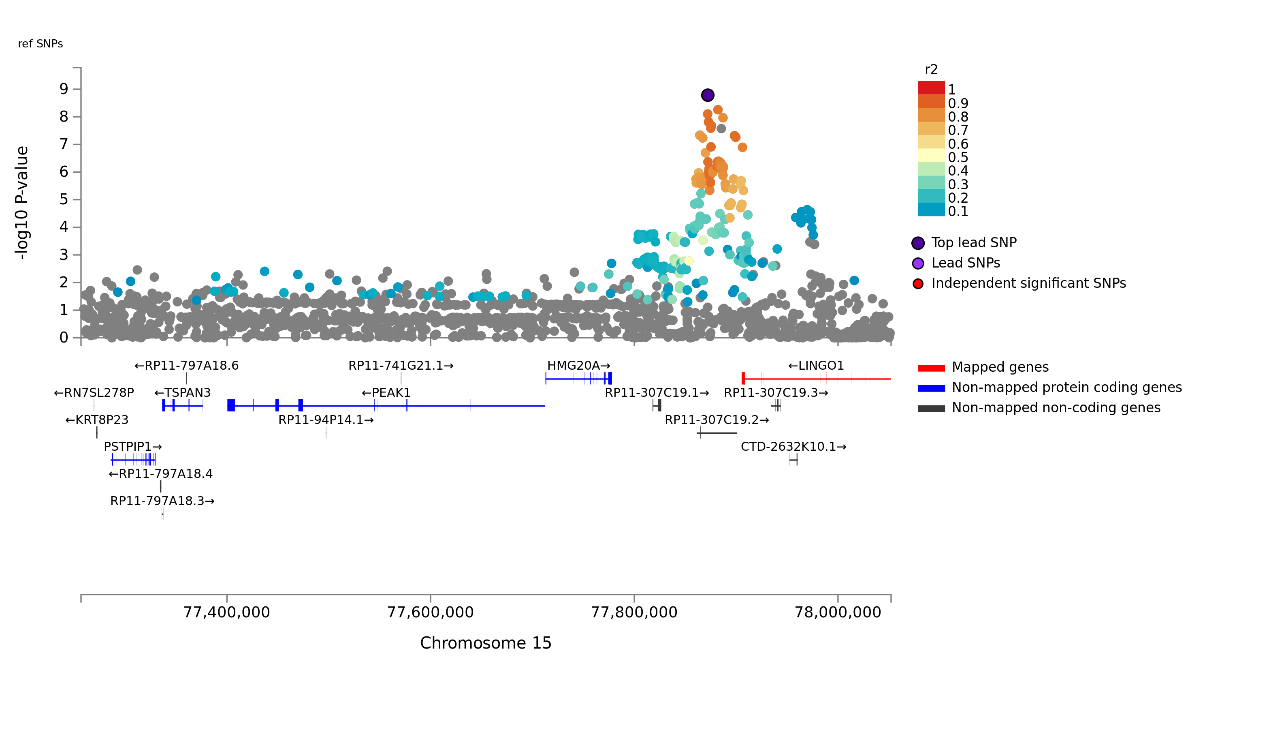

n. locus 14

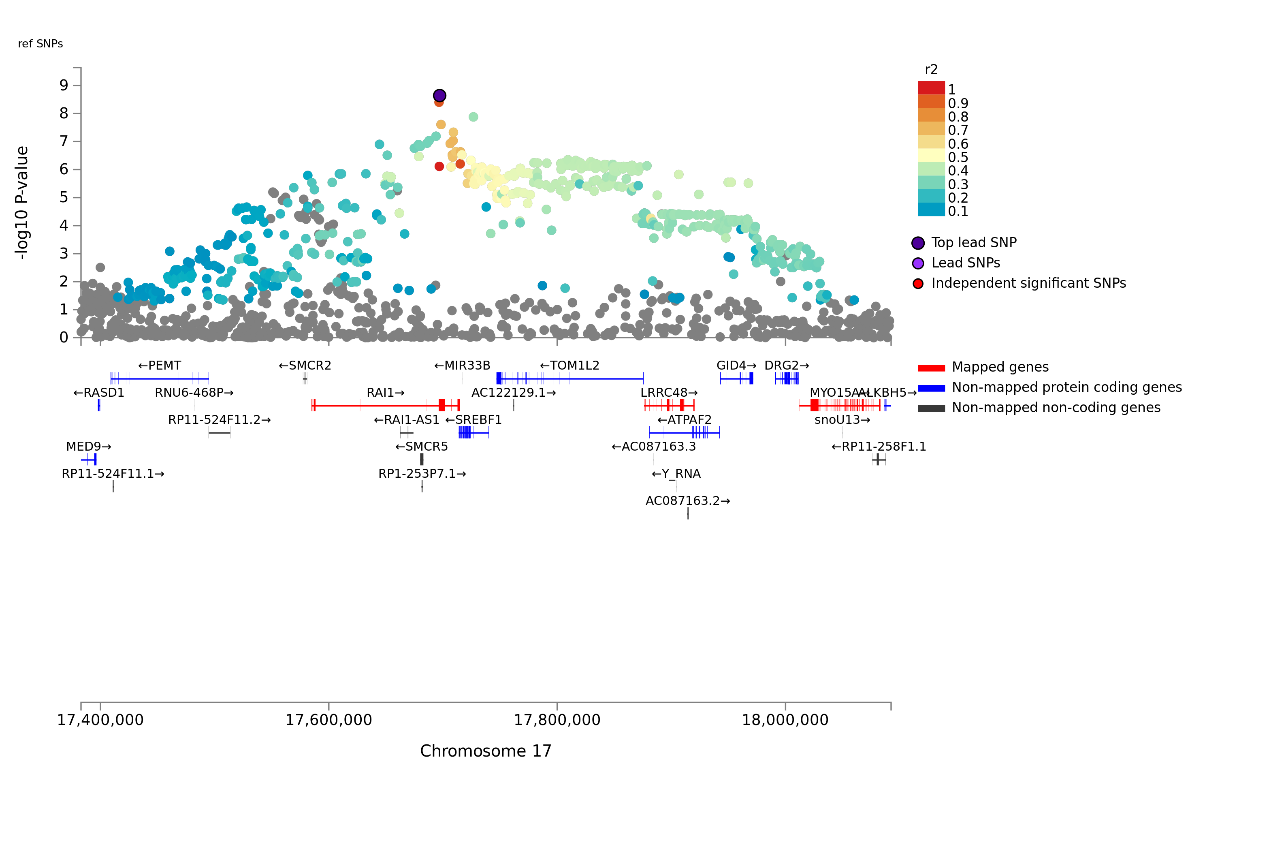

o. locus 15

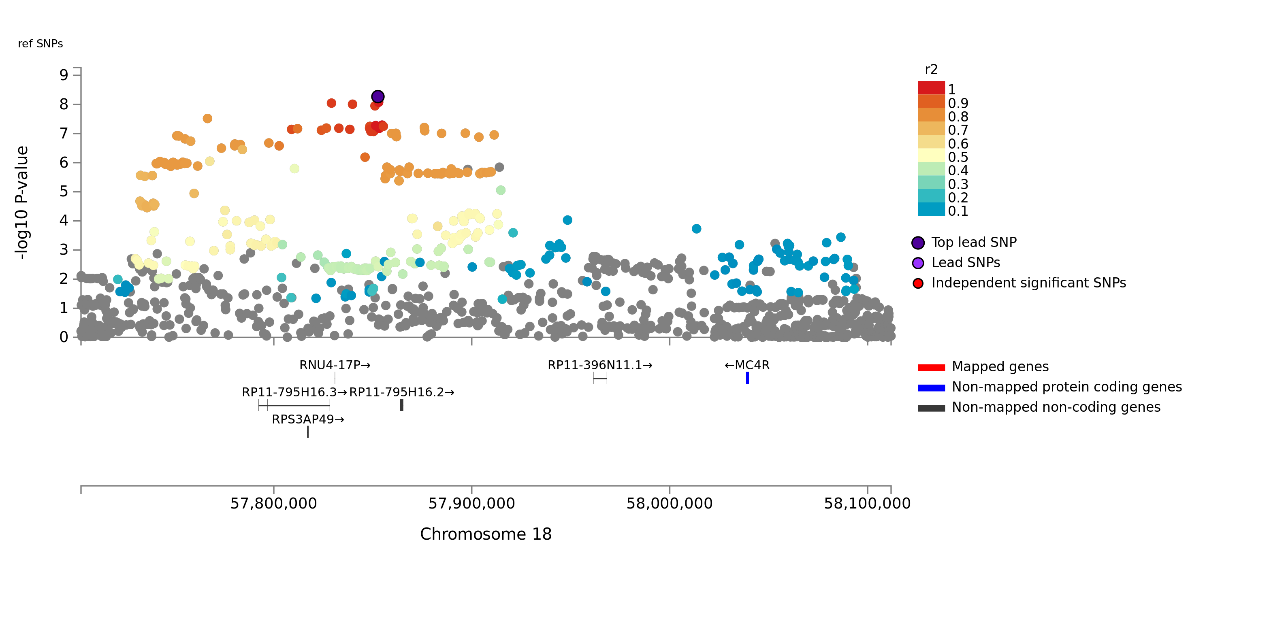

p. locus 16

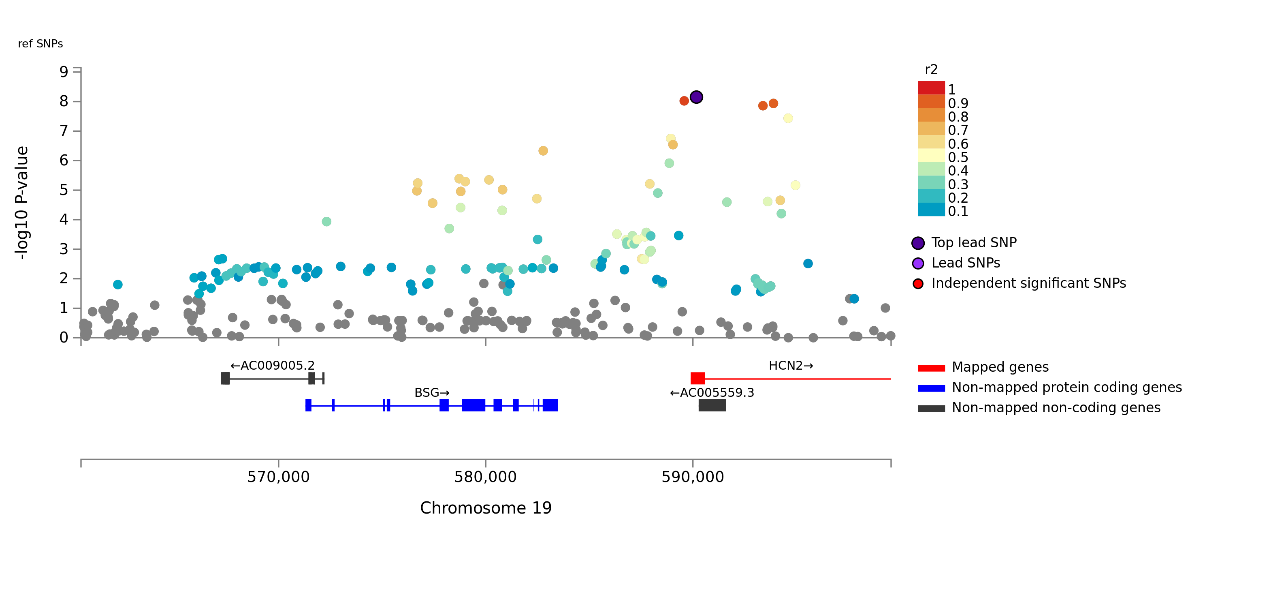

q. locus 17

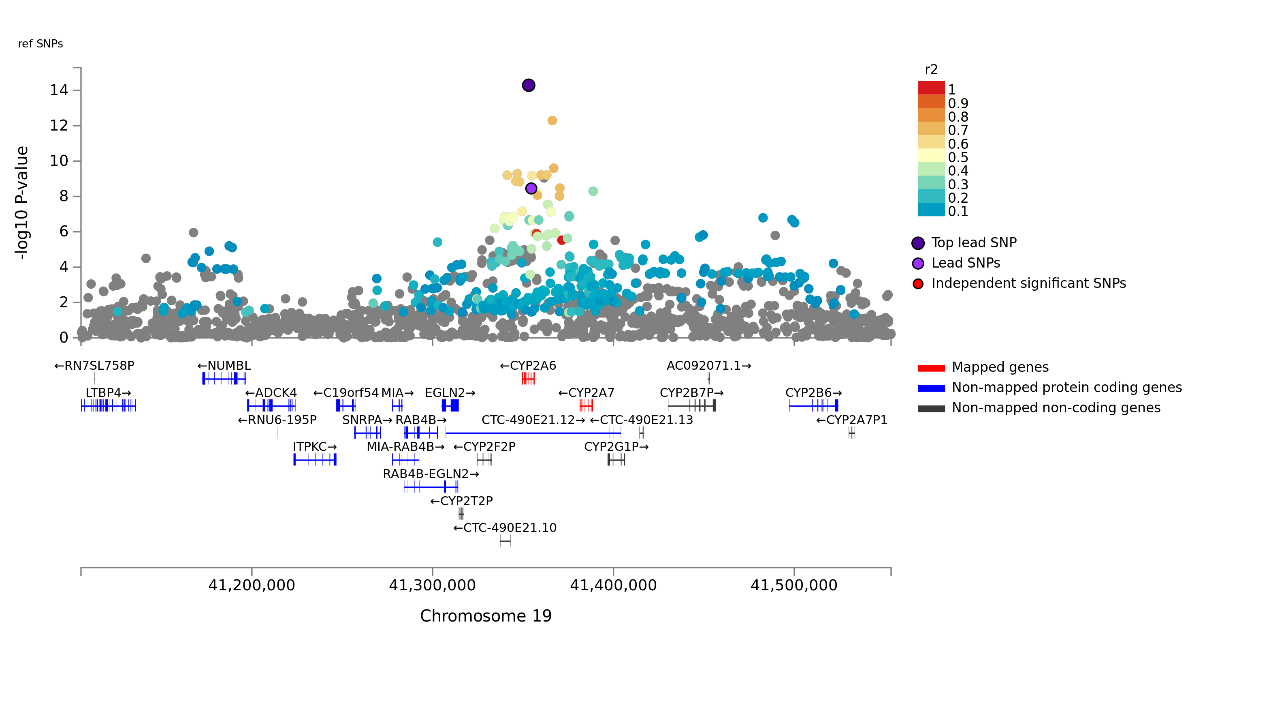
r. locus 18

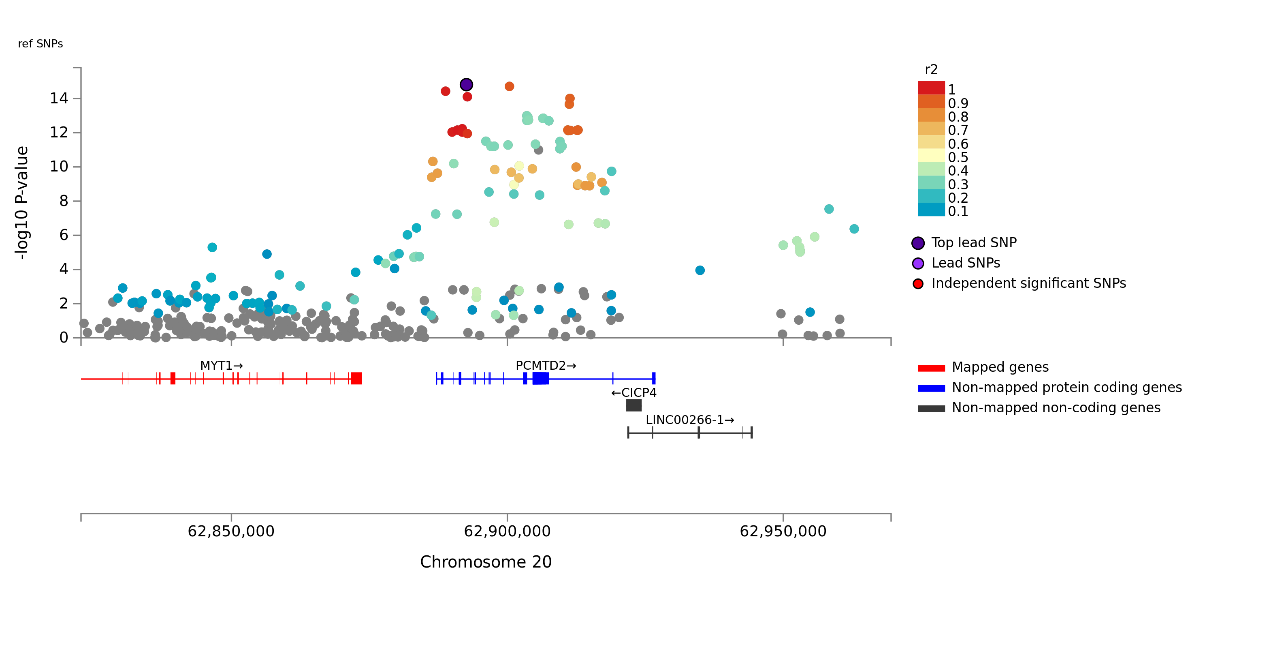

s. locus 19

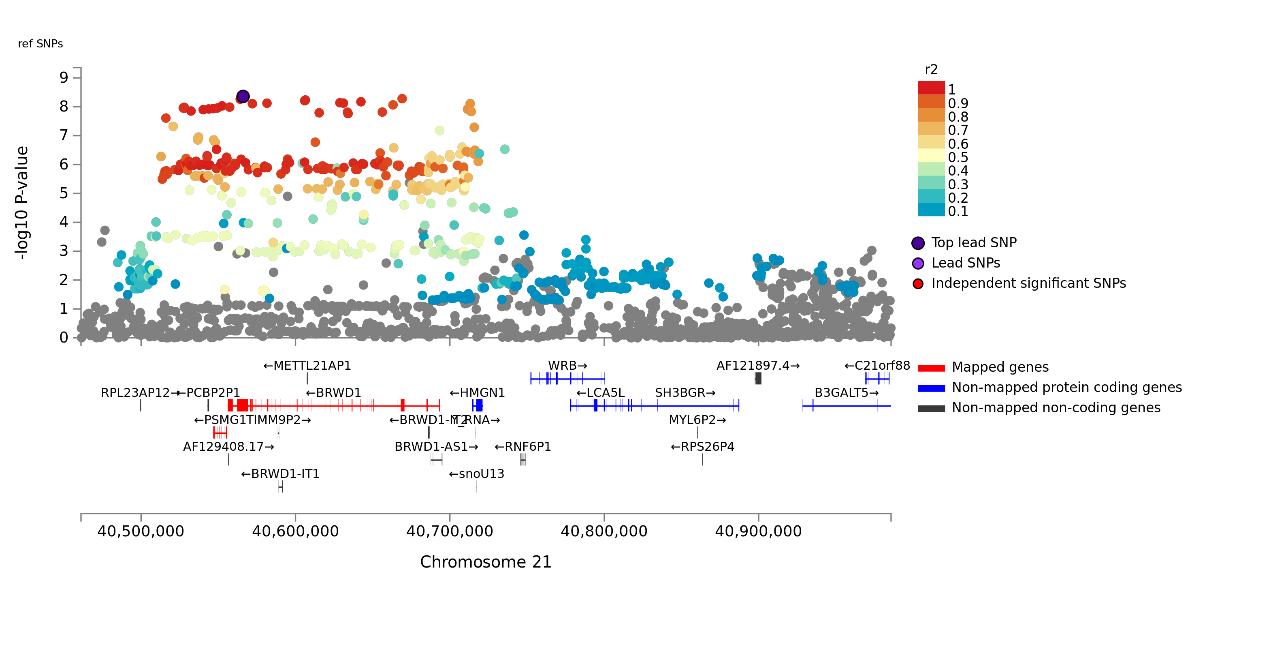

**Supplementary Figure 3. QQ plot for genome-wide analysis study of caffeine consumption among 283,926 coffee consumers. The QQ plot displays the expected –log10 (p-values) on the x-axis and the observed –log10 (p-values) on the y-axis.**
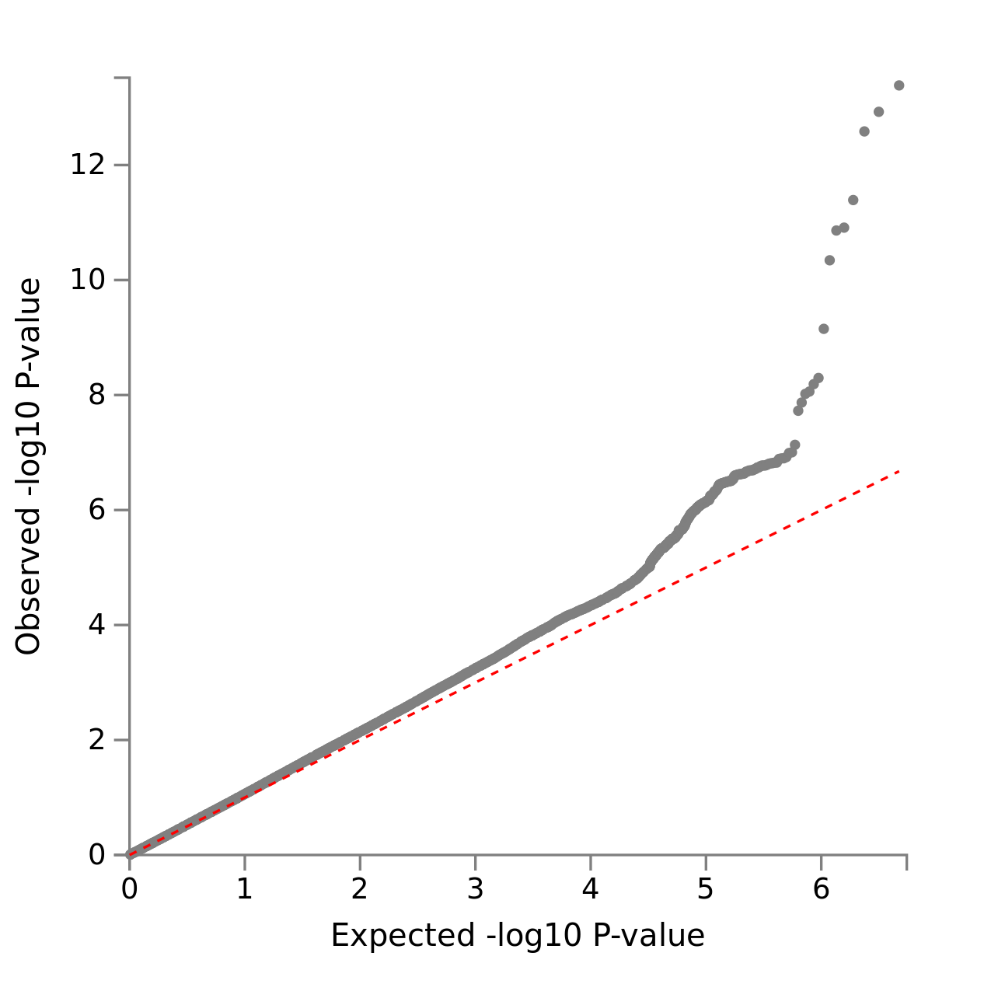

**Supplementary Figure 4. Regional plot for the 2 significant loci of caffeine consumption. Each plot represents a SNP, which are color-coded based on the highest r2 to one of the leading SNPs. SNPs with r2 < 0.05 are colored in grey. The red colored gene were mapped by FUMA using positional mapping.**

**a. locus 1**

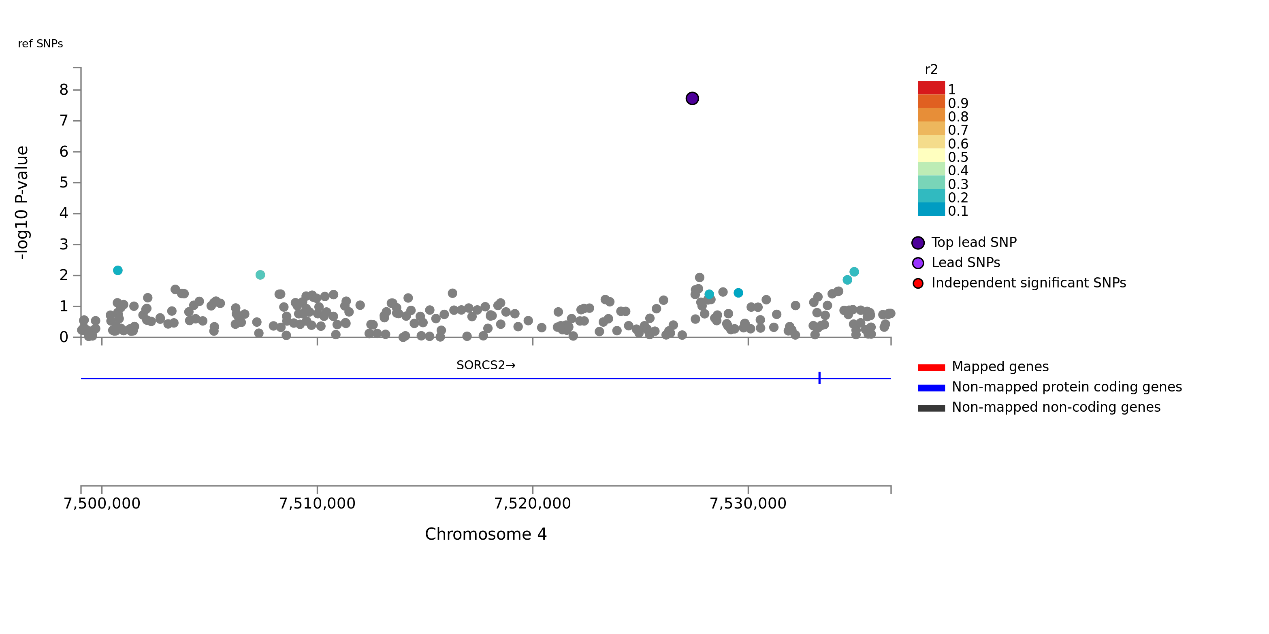

**b. locus 2**

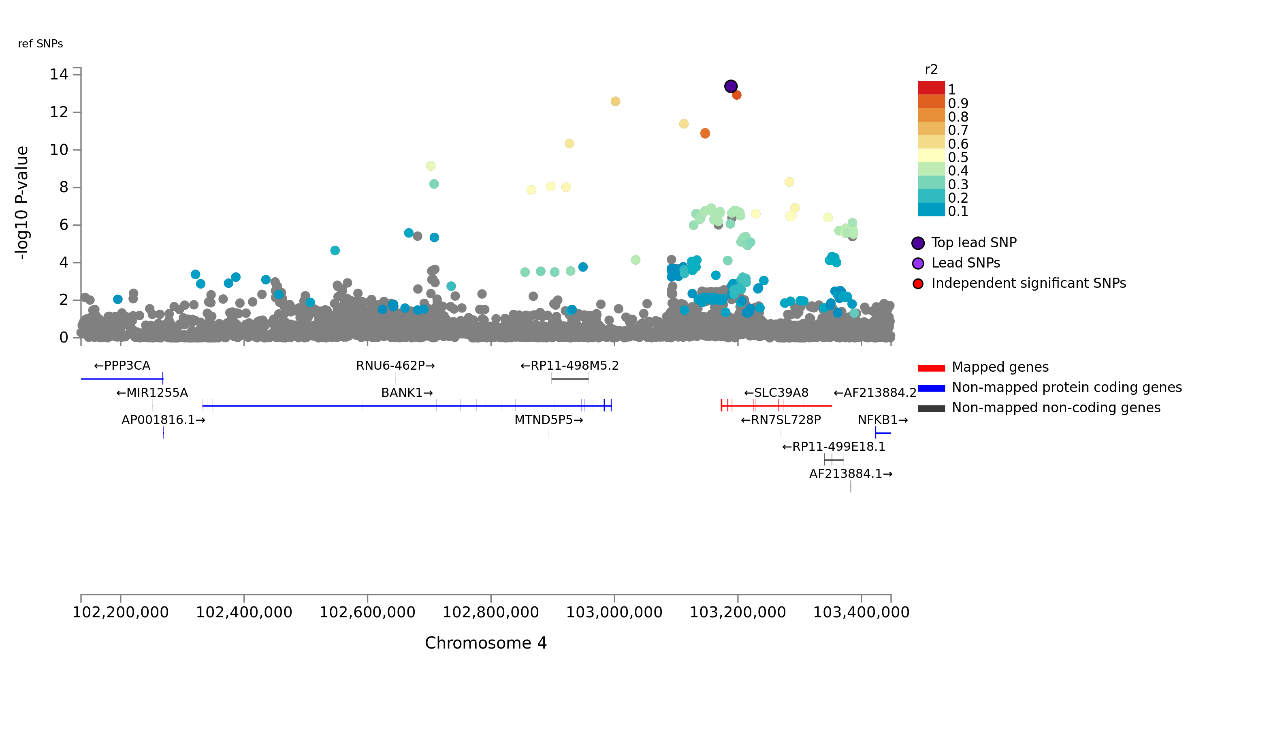

**Supplementary Figure 5. QQ plot for genome-wide analysis study of other non-caffeine substances contained in coffee among 137,371 participants. The QQ plot displays the expected –log10 (p-values) on the x-axis and the observed –log10 (p-values) on the y-axis.**

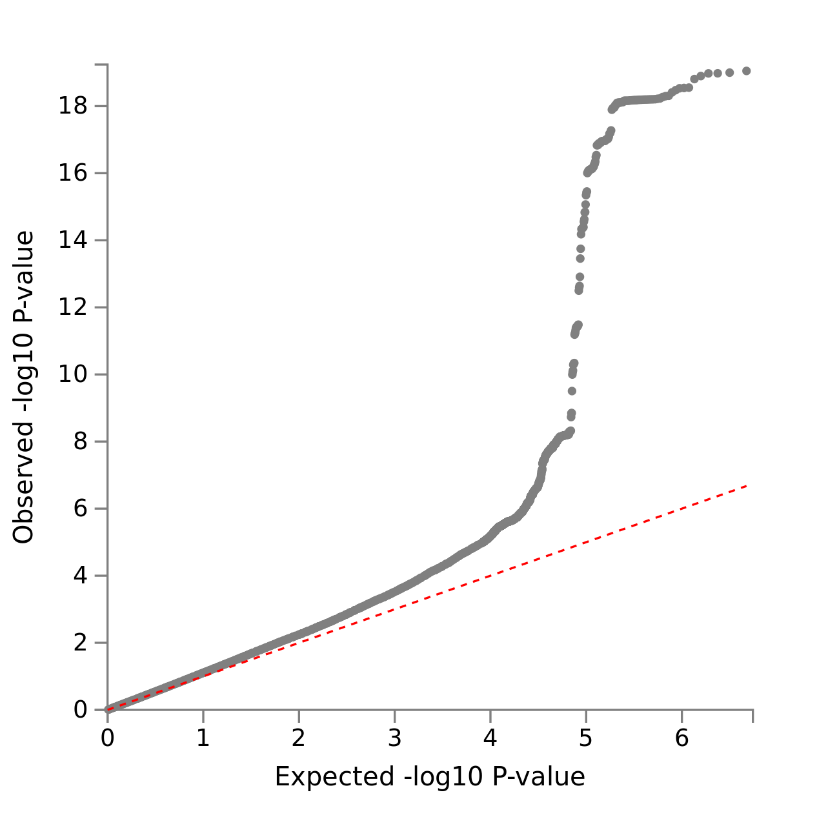

**Supplementary Figure 6. Regional plot for the 5 significant loci of other non-caffeine substances contained in coffee. Each plot represents a SNP, which are color-coded based on the highest r2 to one of the leading SNPs. SNPs with r2 < 0.05 are coloured in grey. The red coloured gene were mapped by FUMA using positional mapping.**

**a. locus 1**

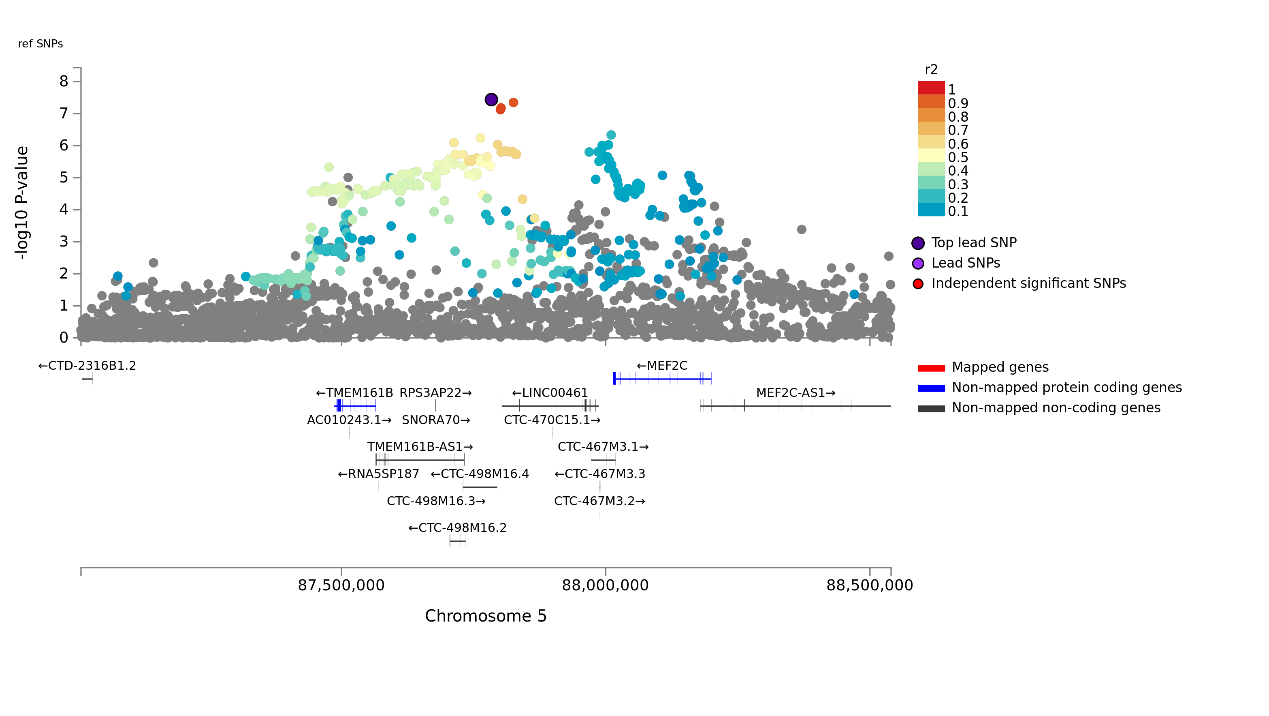

**b. locus 2**

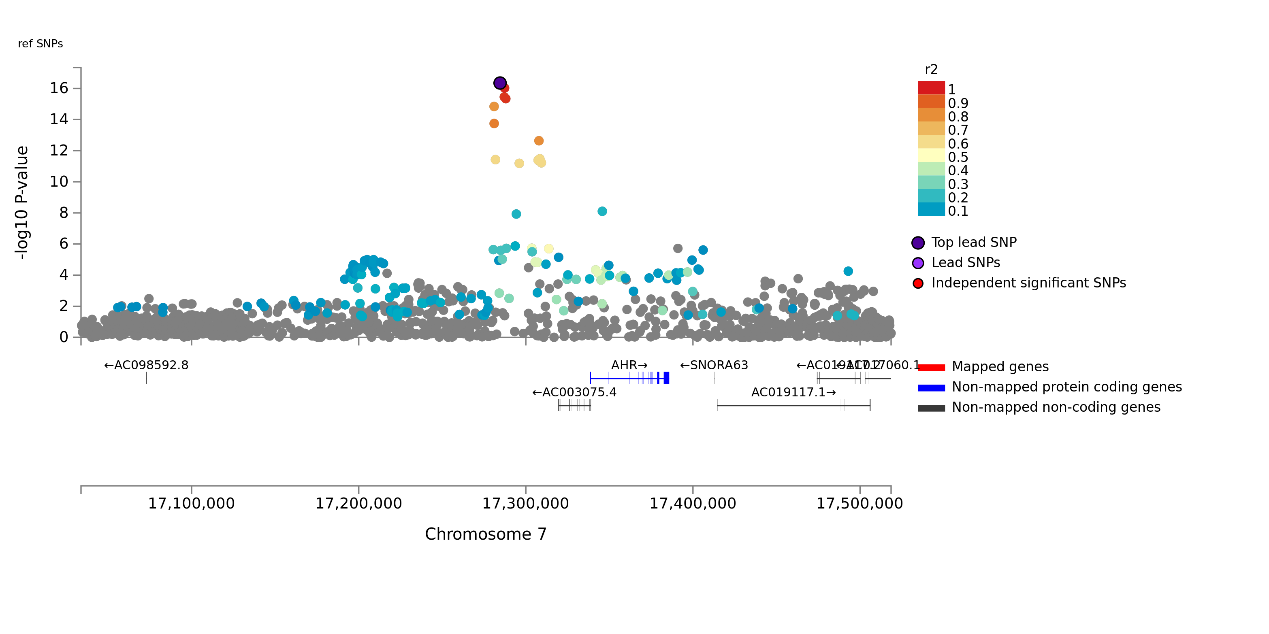

**c. locus 3**

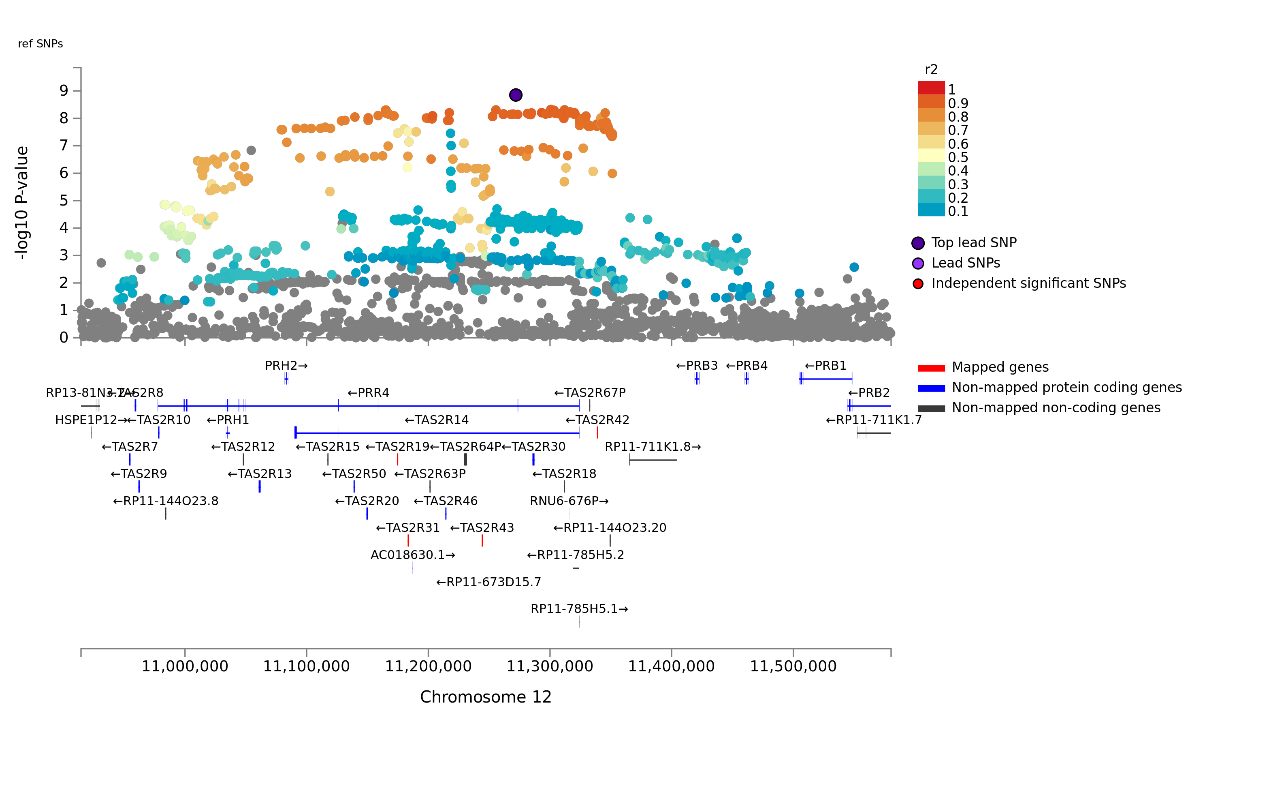

**d. locus 4**

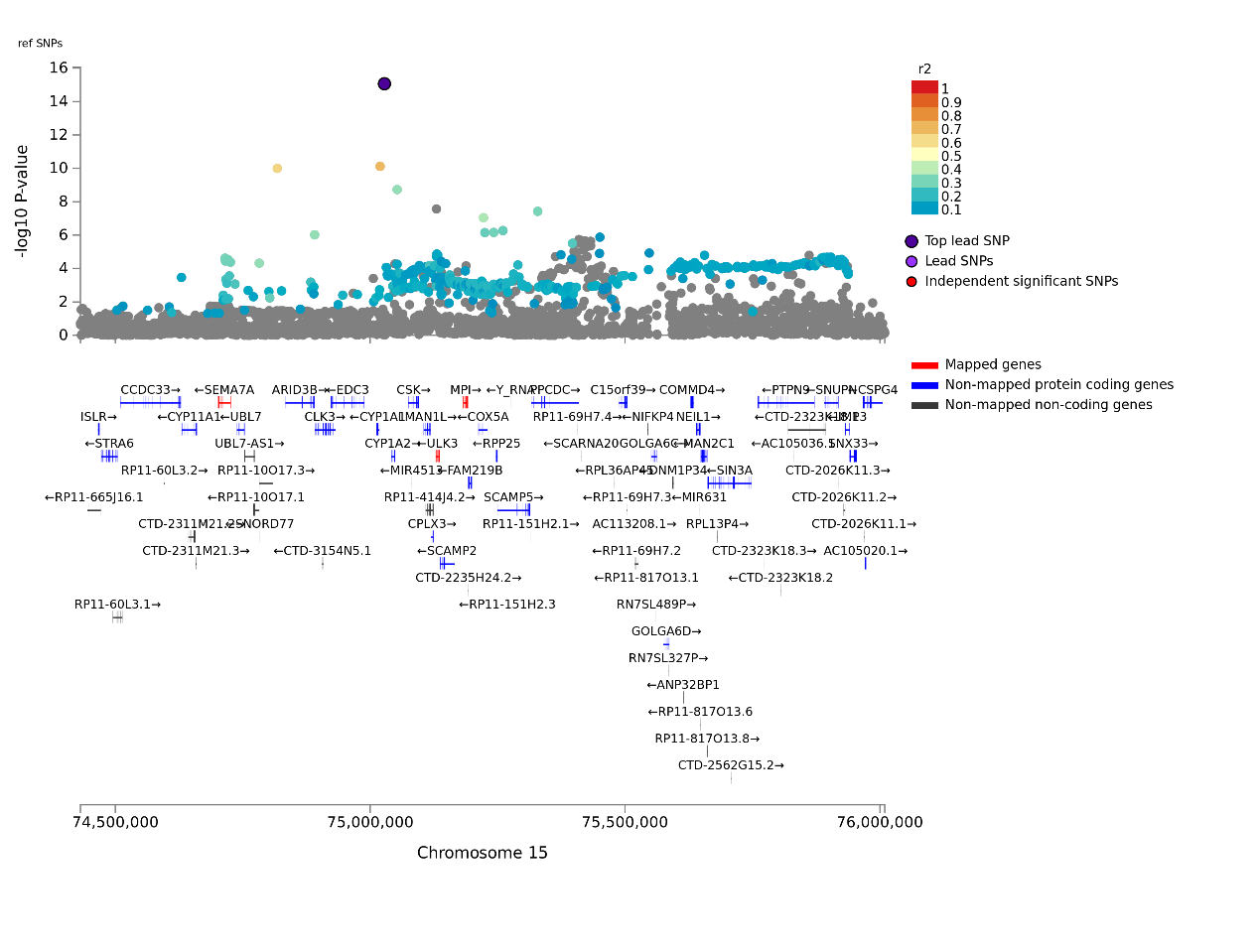

**e. locus 5**

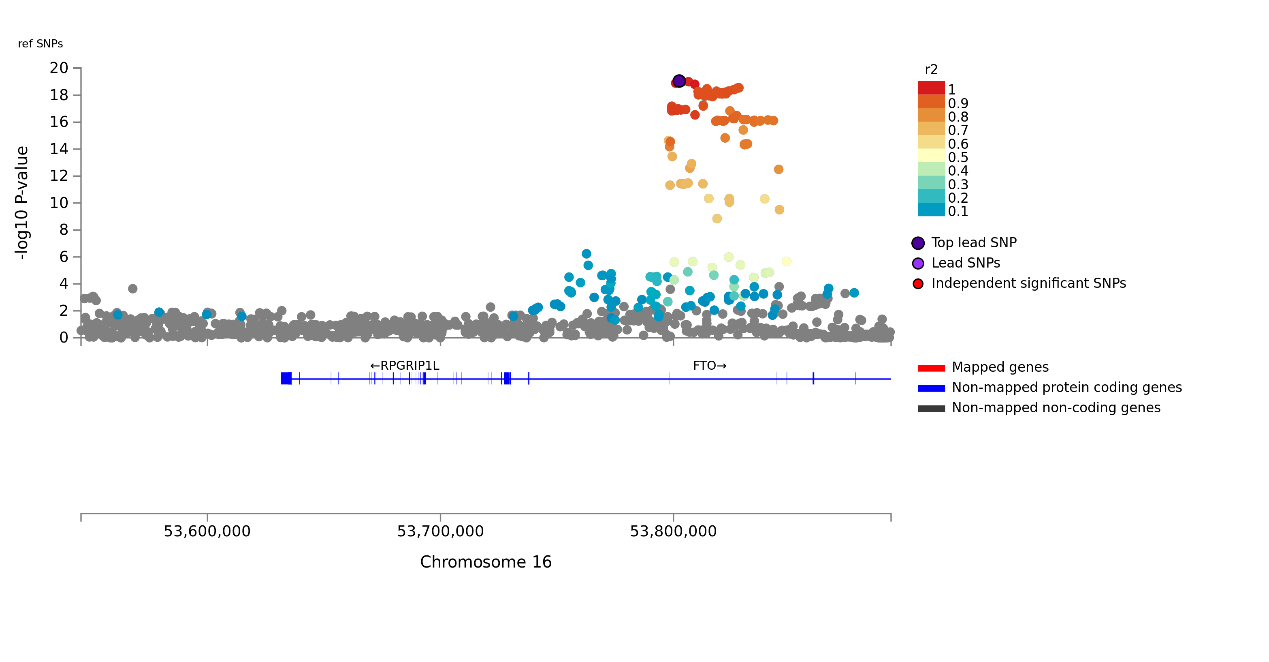

**Supplementary Figure 7.** Functional consequences of SNPs associated with coffee consumption on genes

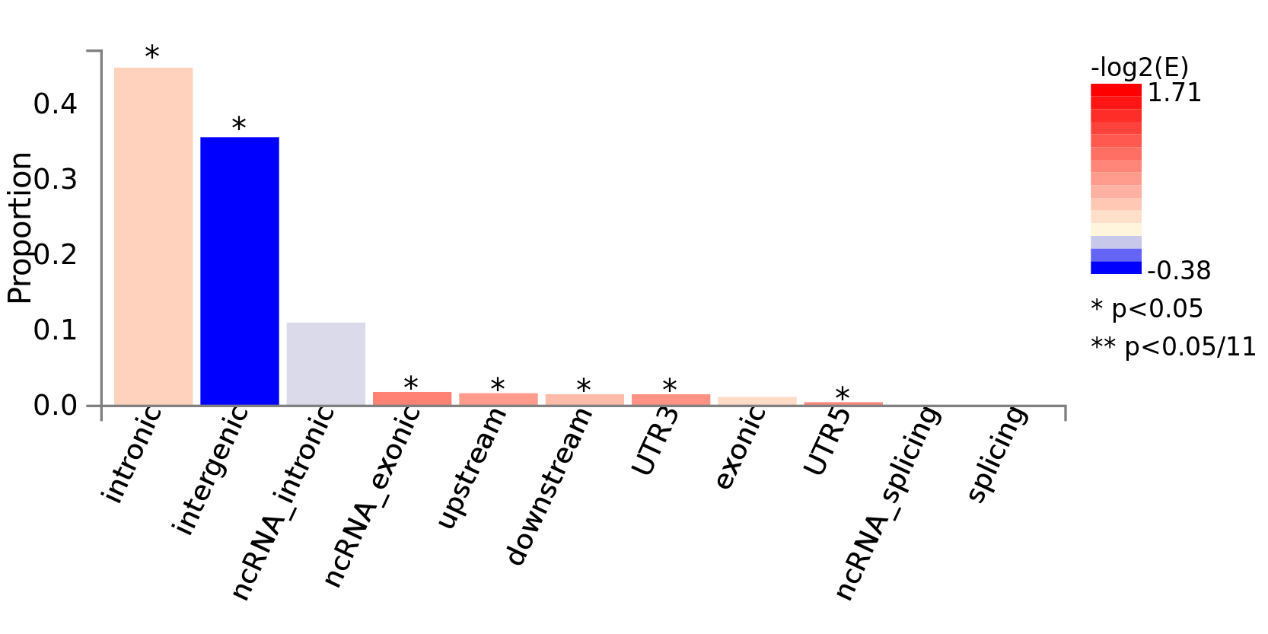

**Supplementary Figure 8.** Functional consequences of SNPs associated with caffeine consumption on genes

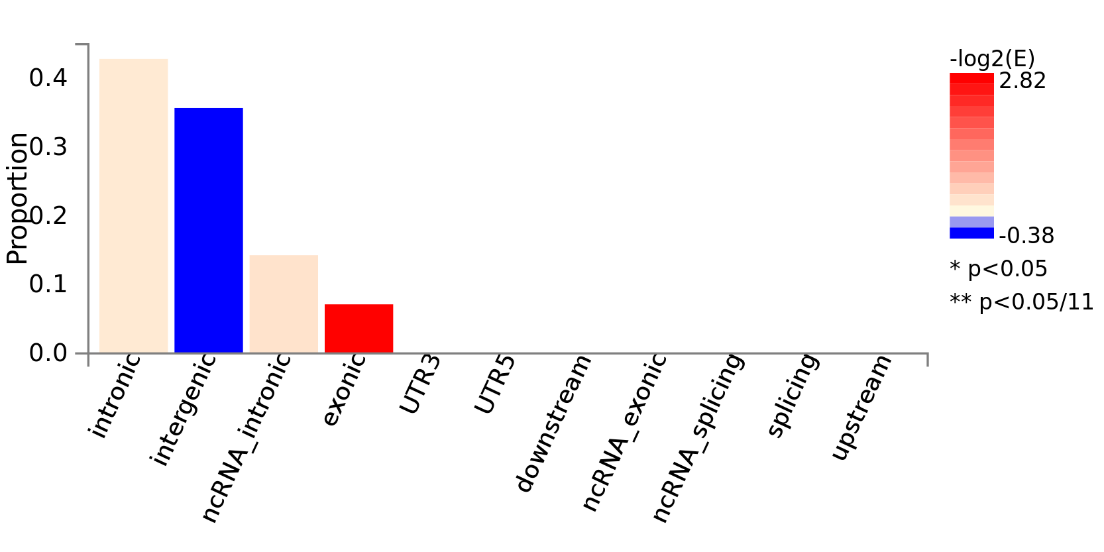
**Supplementary Figure 9.** Functional consequences of SNPs associated with other non-caffeine substances contained in coffee on genes

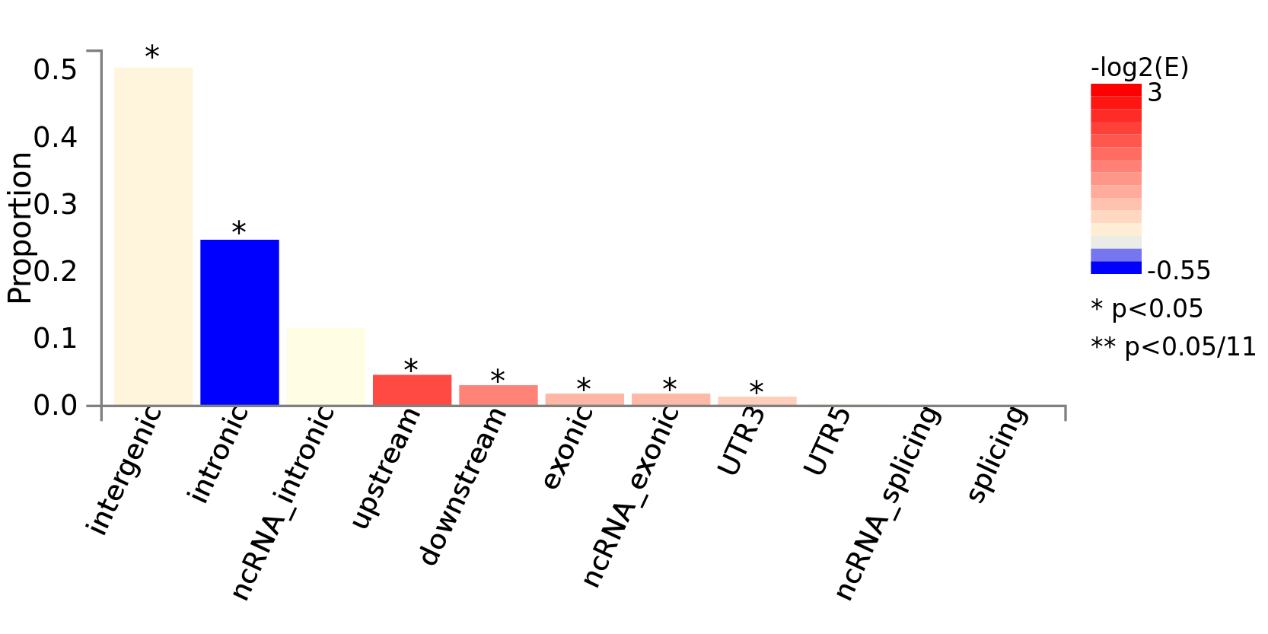
